# Clinical and genetic association studies reveal within-individual variability as an integral part of Huntington disease

**DOI:** 10.1101/2024.05.20.24307629

**Authors:** N. Ahmad Aziz

## Abstract

Huntington disease (HD) is one of the most common repeat expansion disorders. Clinically, HD patients exhibit considerable visit-to-visit variability in symptoms and signs independent of measuring method or rater, yet neither the precise nature nor the determinants of this clinical variability are known. Leveraging detailed genetic and longitudinal clinical data from large cohorts of HD patients, this work *1)* establishes within-individual variability in disease expression as an integral part of HD semiology, demonstrating that it increases with disease duration, mutation size, younger age-at-onset, and lower body weight, *2)* provides a mathematical framework linking higher phenotypic variability to increased predictive entropy, revealing a fundamental relation between within-individual variability and energy expenditure, and *3)* identifies novel genetic modifiers of both within-individual variability and age-at-onset in HD. Thus, accounting for within-individual variability in HD could facilitate the discovery of pathogenic mechanisms and outcomes directly relevant to the development of disease modifying therapies.

## Introduction

Huntington disease (HD) is an autosomal dominantly inherited neurodegenerative disorder caused by a cytosine-adenine-guanine (CAG) repeat expansion in the first exon of the huntingtin (*HTT*) gene. Involuntary movements, cognitive decline, psychiatric and behavioural disturbances as well as progressive weight loss characterise the disease.^1, 2^ We and others have previously demonstrated that the CAG repeat size of the mutant allele is the strongest determinant of both age-at-onset and rate of disease progression in HD.^3^ Apart from indicating progressive global multi-domain impairment, clinical rating scores in HD also exhibit substantial visit-to-visit within-individual variability, which is generally assumed to reflect either limitations of the applied rating scales or intra- and interrater variability.^4^ However, seeing and treating HD patients over many years, I have the impression that disease expression itself can exhibit large visit-to-visit variability on both subjective and (semi-)quantitative metrics *independent* of the measuring methods employed or the rater, yet neither the precise nature nor the determinants of this clinical variability have been investigated so far in HD.

Emerging evidence indicates that not only the mean trajectories of (clinical) biomarkers, but also their intraindividual variability in time constitute risk factors for a range of different diseases.^5^ For example, visit-to-visit variability in blood pressure has been associated with an increased risk of mortality, coronary heart disease, stroke, cognitive decline and dementia,^6-9^ while glycemic variability in diabetic patients has been related to an increased risk of cardiovascular complications.^10^ Similarly, increased body weigh variability has been linked to an increased risk of cardiovascular diseases and cognitive decline.^11, 12^ In patients with common age-associated neurodegenerative disorders like Alzheimer disease and Parkinson disease, increased visit-to-visit variability in body weight and blood pressure have been associated with a faster rate of disease progression.^13, 14^ However, whether intraindividual variability in body weight and other clinical metrics is a characteristic of HD and related to mutation size or disease progression is unknown.

Visit-to-visit within-individual variability of disease expression could reflect the breakdown of central homeostatic mechanisms in HD.^3^ In this scenario, larger mutation size could not only be expected to result in a younger age-at-onset and a faster rate of disease progression, but also in a larger degree of visit-to-visit variability in measures of disease expression, reflecting more extensive underlying homeostatic dysregulation. Therefore, leveraging extensive longitudinal data from the largest cohort study of HD patients, as well as recent advances in the statistical modeling of within-individual variability trajectories across time, this work aims to: *1)* quantify both the extent and characteristics of within-individual variability in clinical measures of disease expression in HD, *2)* assess whether mutation size is associated with longitudinal within-individual clinical variability, *3)* provide a Bayesian brain theory-based mathematical framework in which within-individual variability could be understood as a direct consequence of increased predictive entropy due to progressive neuropathology, *4)* elucidate the genetic determinants of within-individual variability in HD, and *5)* illustrate how treating variability as a core feature of HD could provide insights into the pathogenesis of weight loss, as well as facilitate the discovery of novel genetic modifiers of age-at-onset. The findings indicate that within-individual variability in disease expression is indeed an integral part of HD semiology, with the extent of this variability increasing with both disease duration and mutation size. Moreover, genome-wide association studies (GWAS) identified several genetic modifiers of within-individual variability, indicating that investigating this thus far hidden but integral feature of HD could facilitate the identification of new pathogenic mechanisms with direct relevance for the development of disease modifying therapies for this devastating disorder.

## Methods

### Study Cohort

Monitored data from the Enroll-HD study were used, which also included longitudinal data on a subset of individuals who had previously participated in the Registry study.^15^ Enroll-HD is a global clinical research platform designed to facilitate clinical research in HD. All sites are required to obtain and maintain local ethics committee approvals. Further details are available on the study’s website: enroll-hd.org. All data from the Enroll-HD website were retrieved on 24 December 2020, and included all participants with a mutant *HTT* CAG repeat size between 36 and 65 (to exclude allele counts < 10), body mass index (BMI) < 50 kg/m^2^, and at least three study visits, resulting in an analytical sample size of 7583 individuals (54% female).

### Clinical Measurements

For assessing disease expression, apart from BMI, the Unified Huntington’s Diseases Rating Scale (UHDRS) total motor score, total functional capacity (TFC), scores on the Symbol Digit Modality Test (SDMT) and Stroop Word Reading Test (SWRT), as well as a composite clinical outcome measure derived from these individual UHDRS domain scores as described previously,^4^ were used. Except for the UHDRS total motor score, higher scores indicate better clinical performance.

### Genotype data

Individual-level imputed genotype data from the largest GWAS of residual age-at-onset in HD were obtained from the Genetic Modifiers of Huntington Disease (GeM-HD) Consortium.^16^ This GWAS included data on a total of 9064 HD patients. Residual age-at-onset was defined as the difference in years between observed and expected age of motor onset based on mutant CAG repeat size.^3, 16^ Individual-level imputed genotype data were available for a subset of 2230 HD patients from the Enroll-HD study, 1312 of whom were also included in the Gem-HD GWAS. Details of the genotyping and imputation procedures have been reported previously.^16^

*Genome-wide association study of mean and within-individual variability of clinical measures* In the subset of individuals from Enroll-HD in whom genotype data were available, two separate GWAS studies were conducted to identify the genetic modifiers of both mean and within-individual variability in the UHDRS total motor score, SDMT score, and BMI. Only participants with at least three study visits and a mutant *HTT* CAG repeat size between 38 and 52 (in order to exclude allele counts < 10) were included. Additionally, extensive per individual and per variant quality control (QC) were performed using plinkQC,^17^ applying the following (default) filters for inclusion: genotyping rate > 0.97, heterozygosity rate < 3 standard deviations from the sample mean, proportion of identity-by-descent of < 0.1875 for pairs of individuals, European descent (defined as < 3 times the maximum Euclidean distance from the center of the HapMap European reference samples), variant missing rate < 0.01, Hardy-Weinberg equilibrium (p ≥ 1E-5), minor allele frequency ≥ 0.05 and imputation score ≥ 0.5. After filtering and QC, 1292 individuals and 6,625,212 variants remained for analysis.

For the within-individual variability GWAS analyses two different methods were employed. Initially, TrajGWAS, a modified linear mixed effects model-based approach for assessing genetic effects on both the mean and within-individual variability of continuous outcomes^5^ (as implemented in the TrajGWAS.jl Julia package, version 0.4.3, Julia version 1.8.5), was used. A per single-nucleotide polymorphism (SNP) analysis was conducted, using the default parameters and 10 runs with an additive model and a score test with a saddle-point approximation for estimating effect sizes and p-values for each genetic variant. The models included age, mutant CAG repeat size, an interaction term between age and mutant CAG repeat size (to adjust for the effect of CAG repeat size on the rate of disease progression), sex, and the first 10 genetic principle components (to account for population stratification) as fixed effects, a random intercept and slope for age (to account for repeated intraindividual measurements), and an intercept only for estimating the remaining within-subject variance. However, given that the TrajGWAS-based estimates of within-individual variability effects were unstable and showed a high degree of genomic inflation (λ ≥ 1.3), these results are not reported here. Instead, a more conservative method was employed for estimating the genetic determinants of within-individual variability in HD, as follows: *1)* using the entire analytical Enroll-HD dataset (N=7583), a random intercept and slope linear mixed effects model was fitted with sex, age, age squared, mutant CAG repeat size, and an interaction term between age and mutant CAG repeat size as fixed effects, and age as a random effect, and the clinical measures as the dependent variables, *2)* the remaining within individual variability was extracted as the mean of the sum of the squared errors around the fitted random regression lines for each outcome measure, *3)* a logarithmic transformation (given the right-skewed distribution) and scaling to a standard normal distribution was performed, and *4)* these estimates were used as the dependent variable in a mixed-effects model GWAS employing Genome-wide Efficient Mixed Model Association (GEMMA, version 0.98.5).^18^ Briefly, in GEMMA, within-individual variability estimates were modeled as a function of minor allele dosage, sex, mutant CAG repeat size, and the first four genetic principle components, while also adjusting for remaining population and sample structure in a univariate linear mixed effects model as previously applied by the Gem-HD consortium.^16^ A similar approach was used to perform a GWAS on the means of the clinical measures, except now employing a random intercept only linear mixed effects model to estimate the mean levels of each clinical outcome measure (adjusted for sex, age, age squared, CAG repeat size, and its interaction with age) for each individual during the follow-up period.

### Genome-wide association study of age of onset

To account for mutant CAG repeat size-dependent increases in phenotypic variability, a GWAS of *relative* residual age-at-onset (defined as residual age-at-onset divided by the expected age of onset) was performed using an otherwise identical statistical model and the same dataset (N = 9064) as previously reported by the Gem-HD consortium.^16^

### Genomic risk loci and gene-based analysis

Functional Mapping and Analysis of GWAS (FUMA) was used to identify genomic risk loci.^19^ Genome-wide significant SNPs in relatively high linkage disequilibrium (LD) (i.e., r^2^ > 0.6) with nearby SNPs were used to define genomic risk loci. Variant annotation for each locus was based on lead SNPs and candidate SNPs, defined as those SNPs in LD with the lead SNP within a window of 250 kb and nominally significant at p < 0.05. Functionally annotated SNPs were subsequently mapped to genes based on physical position (positional mapping) and expression quantitative trait loci (eQTL mapping). MAGMA was used to conduct gene and gene-set analysis,^20^ summarizing SNP-trait associations at the level of genes and gene sets and mapping these to biological pathways, using a Bonferroni-correction for multiple testing. This was followed by tissue enrichment analysis to investigate tissue specificity using the GTEx database (version 8). For pathway enrichment analysis, mapped genes were further investigated using the GENE2FUNC tool in FUMA,^21^ applying the Benjamini-Hochberg false discovery rate (FDR) method for multiple comparisons correction.

### Statistical Analysis

To assess the association of disease duration and *HTT* CAG repeat size with visit-to-visit clinical variability a recently developed regression method was utilized, i.e., within-subject variance estimator by robust regression (WiSER) (as implemented in the WiSER.jl Julia package, version 0.2.3), which is specifically designed to assess the effects of both time-dependent and time-independent predictors on longitudinal within-individual variability.^22^ WiSER is robust against misspecifications of the distributions of the conditional outcomes and random effects.^22^ As outcomes we used the composite score, as well as its constituents (including the UHDRS total motor score, TFC, and SDMT and SWRT score tests), and BMI. As predictors, age at the time of each visit, a squared term for age to account for non-linear progression, mutant *HTT* CAG repeat size, an interaction term between age and mutant *HTT* CAG repeat size, and sex were included, applying a random intercept and slope model to account for repeated measurements. Age was used as the time indicator to allow for simultaneous adjustment for possible age effects on disease progression, which would not have been possible with inclusion of disease duration due to a high degree of collinearity between age and disease duration.^3^ To assess the association between within-individual variability and age-at-onset, residual age-at-onset was added as an independent variable to the previous model. The robustness of the findings was assessed in two additional sensitivity analyses. In the first sensitivity analysis, the same approach as detailed in the previous section (*‘Genome-wide association study of mean and within-individual variability of clinical measures’*) was used for calculating the individual level variability as the mean of the sum of the squared errors around the fitted random intercept and slope regression lines for each outcome measure, comparing the values across different mutant CAG repeat size categories using the non-parametric Wilcoxon’s rank sum test. In a second sensitivity analysis, the variation independent of mean (VIM) metric was calculated as described previously.^9^ For calculating the VIM of composite score, first a constant of 10 was added to offset the composite scores as VIM is undefined over negative values.^9^ After regressing out the effects of sex, age at baseline, and domain-specific disease severity at baseline (e.g., for motor score, the motor score at baseline was used as a measure of disease severity) from the VIM scores, the residuals were compared across different categories of mutant CAG repeat size using the non-parametric Wilcoxon’s rank sum test. A similar approach was employed to visualize the association between residual age-at-onset and VIM of the different outcomes, where an additional adjustment for mutant CAG repeat size and its interaction with age at baseline were included. Programming was performed in R (base version 4.2.1) and Julia (version 1.8.5). Two-tailed p-values < 0.05 were considered statistically significant.

## Results

### Part 1: Within individual variability is an integral feature of HD

The baseline characteristics of the study sample are included in **Supplementary Table 1**. As expected, age and CAG repeat size, as well as their interaction, were highly significantly association with disease progression across all six outcome measures. On average, the TFC score and BMI were higher, and the SDMT score was lower, in men compared to women (**Table 1**).

**Table 1.**
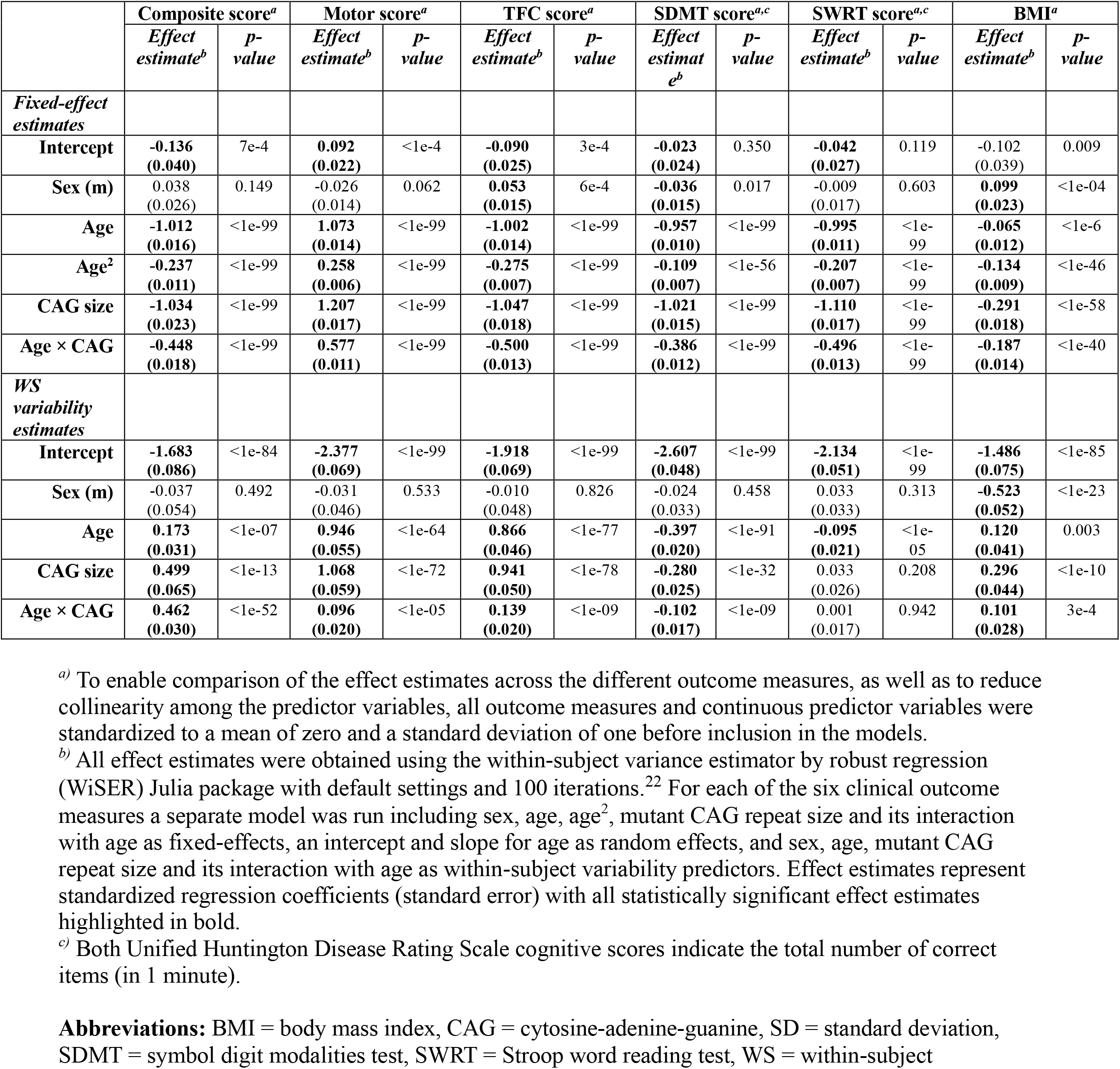
Summary of model estimates.

|  | Composite score<sup>a</sup> |  | Motor score<sup>a</sup> |  | TFC score<sup>a</sup> |  | SDMT score<sup>a,c</sup> |  | SWRT score<sup>a,c</sup> |  | BMI<sup>a</sup> |  |
| --- | --- | --- | --- | --- | --- | --- | --- | --- | --- | --- | --- | --- |
|  | Effect estimate<sup>b</sup> | p-value | Effect estimate<sup>b</sup> | p-value | Effect estimate<sup>b</sup> | p-value | Effect estimate<sup>b</sup> | p-value | Effect estimate<sup>b</sup> | p-value | Effect estimate<sup>b</sup> | p-value |
| *Fixed-effect estimates* |  |  |  |  |  |  |  |  |  |  |  |  |
| Intercept | -0.136<br>(0.040) | 7e-4 | **0.092**<br>(0.022) | <1e-4 | -0.090<br>(0.025) | 3e-4 | -0.023<br>(0.024) | 0.350 | -0.042<br>(0.027) | 0.119 | -0.102<br>(0.039) | 0.009 |
| Sex (m) | 0.038<br>(0.026) | 0.149 | -0.026<br>(0.014) | 0.062 | **0.053**<br>(0.015) | 6e-4 | -0.036<br>(0.015) | 0.017 | -0.009<br>(0.017) | 0.603 | **0.099**<br>(0.023) | <1e-04 |
| Age | -1.012<br>(0.016) | <1e-99 | **1.073**<br>(0.014) | <1e-99 | -1.002<br>(0.014) | <1e-99 | -0.957<br>(0.010) | <1e-99 | -0.995<br>(0.011) | <1e-99 | -0.065<br>(0.012) | <1e-6 |
| Age<sup>2</sup> | -0.237<br>(0.011) | <1e-99 | **0.258**<br>(0.006) | <1e-99 | -0.275<br>(0.007) | <1e-99 | -0.109<br>(0.007) | <1e-56 | -0.207<br>(0.007) | <1e-99 | -0.134<br>(0.009) | <1e-46 |
| CAG size | -1.034<br>(0.023) | <1e-99 | **1.207**<br>(0.017) | <1e-99 | -1.047<br>(0.018) | <1e-99 | -1.021<br>(0.015) | <1e-99 | -1.110<br>(0.017) | <1e-99 | -0.291<br>(0.018) | <1e-58 |
| Age × CAG | -0.448<br>(0.018) | <1e-99 | **0.577**<br>(0.011) | <1e-99 | -0.500<br>(0.013) | <1e-99 | -0.386<br>(0.012) | <1e-99 | -0.496<br>(0.013) | <1e-99 | -0.187<br>(0.014) | <1e-40 |
| *WS variability estimates* |  |  |  |  |  |  |  |  |  |  |  |  |
| Intercept | -1.683<br>(0.086) | <1e-84 | -2.377<br>(0.069) | <1e-99 | -1.918<br>(0.069) | <1e-99 | -2.607<br>(0.048) | <1e-99 | -2.134<br>(0.051) | <1e-99 | -1.486<br>(0.075) | <1e-85 |
| Sex (m) | -0.037<br>(0.054) | 0.492 | -0.031<br>(0.046) | 0.533 | -0.010<br>(0.048) | 0.826 | -0.024<br>(0.033) | 0.458 | 0.033<br>(0.033) | 0.313 | -0.523<br>(0.052) | <1e-23 |
| Age | **0.173**<br>(0.031) | <1e-07 | **0.946**<br>(0.055) | <1e-64 | **0.866**<br>(0.046) | <1e-77 | -0.397<br>(0.020) | <1e-91 | -0.095<br>(0.021) | <1e-05 | **0.120**<br>(0.041) | 0.003 |
| CAG size | **0.499**<br>(0.065) | <1e-13 | **1.068**<br>(0.059) | <1e-72 | **0.941**<br>(0.050) | <1e-78 | -0.280<br>(0.025) | <1e-32 | 0.033<br>(0.026) | 0.208 | **0.296**<br>(0.044) | <1e-10 |
| Age × CAG | **0.462**<br>(0.030) | <1e-52 | **0.096**<br>(0.020) | <1e-05 | **0.139**<br>(0.020) | <1e-09 | -0.102<br>(0.017) | <1e-09 | 0.001<br>(0.017) | 0.942 | **0.101**<br>(0.028) | 3e-4 |

<sup>a)</sup> To enable comparison of the effect estimates across the different outcome measures, as well as to reduce collinearity among the predictor variables, all outcome measures and continuous predictor variables were standardized to a mean of zero and a standard deviation of one before inclusion in the models.
<sup>b)</sup> All effect estimates were obtained using the within-subject variance estimator by robust regression (WiSER) Julia package with default settings and 100 iterations.<sup>22</sup> For each of the six clinical outcome measures a separate model was run including sex, age, age<sup>2</sup>, mutant CAG repeat size and its interaction with age as fixed-effects, an intercept and slope for age as random effects, and sex, age, mutant CAG repeat size and its interaction with age as within-subject variability predictors. Effect estimates represent standardized regression coefficients (standard error) with all statistically significant effect estimates highlighted in bold.
<sup>c)</sup> Both Unified Huntington Disease Rating Scale cognitive scores indicate the total number of correct items (in 1 minute).
**Abbreviations:** BMI = body mass index, CAG = cytosine-adenine-guanine, SD = standard deviation, SDMT = symbol digit modalities test, SWRT = Stroop word reading test, WS = within-subject

#### Within-individual variability increases with disease duration and CAG repeat size

Visit-to-visit within-individual variability in the composite, motor and functional scores and BMI markedly increased with both age and mutant CAG repeat size, whereas cognitive test score variability decreased for SDMT with higher age and CAG repeat size (**Table 1**). Importantly, the effects of age and mutant CAG repeat size on visit-to-visit variability were synergistic across all outcome measures as indicted by highly significant interaction effects with similar directions to the main effects (**Table 1**). Men exhibited lower BMI variability, but otherwise no sex differences in variability of disease expression were observed. Except for cognitive scores, both sensitivity analyses confirmed the findings based on the WiSER method, showing a progressively higher within-individual variability with increasing mutant CAG repeat size for the composite, motor and functional scores, as well as BMI (**Figure 1** and **Supplementary Figure 1**). However, the sensitivity analyses, which employed groupings based on mutant CAG repeat size, indicated that within-individual variability in cognitive scores (SDMT and SWRT scores) also increased with larger mutant CAG repeat size. The difference between the WiSER-based estimates and the mutant CAG repeat size-stratified analyses for cognitive scores could be due to the levelling off of the variability for larger mutation sizes (**Figure 1** and **Supplementary Figure 1**), which may reflect floor/ceiling effects of the cognitive testing batteries in HD.^23^

**Figure 1.**
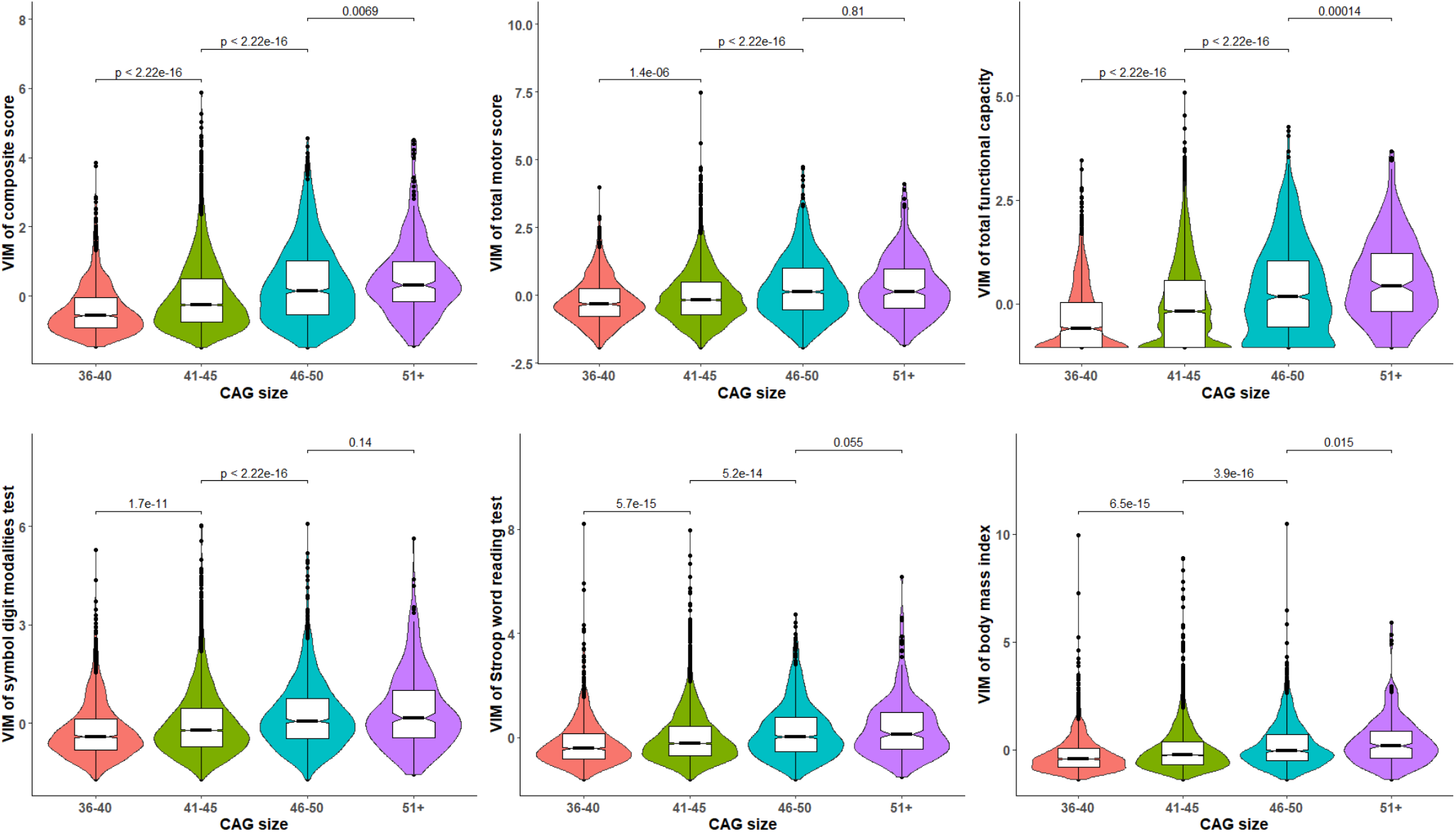
Within-individual clinical variability in Huntington disease increases with larger mutant *HTT* CAG repeat size. The violin plots depict the association of mutant *HTT* CAG repeat size and the standardized variance independent of the mean (VIM) metric for the Unified Huntington Disease Rating Scale composite score, total motor score, total functional capacity, and symbol digit modalities and Stroop word reading test scores, as well as body mass index. VIM of all clinical measures significantly increased with larger mutant *HTT* CAG repeat size, although for the cognitive scores, the effect tended to level off for larger mutation sizes. The boxes indicate the interquartile ranges around the median (thick black lines), with values deviating more than 1.5 times the interquartile range from the median represented by black dots. After regressing out the effects of sex, age at baseline, and domain-specific disease severity at baseline (e.g., for composite score, the composite score at baseline was used as a measure of disease severity) from the VIM scores, the residuals were compared across different categories of mutant CAG repeat size using the non-parametric Wilcoxon’s rank sum test.

#### Within-individual variability increases with younger age-at-onset

A lower residual age-at-onset was robustly associated with higher visit-to-visit within-individual variability in all clinical outcome measures, including the UHDRS composite, motor, functional and cognitive scores, as well as BMI, independent of sex, age or mutation size (**Table 2** and **Figure 2**).

**Table 2.** The association between residual age-at-onset and within-subject variability.

|  | Residual age-at-onset |  |  |  |
| --- | --- | --- | --- | --- |
|  | *Fixed-effect estimates* |  | *WS variability estimates* |  |
|  | *Effect estimate<sup>b</sup>* | *p-value* | *Effect estimate<sup>b</sup>* | *p-value* |
| **Composite score<sup>a</sup>** | **0.595 (0.016)** | <1e-99 | **-0.057 (0.029)** | 0.046 |
| **Motor score<sup>a</sup>** | **-0.483 (0.026)** | <1e-78 | **-0.707 (0.065)** | <1e-27 |
| **TFC score<sup>a</sup>** | **0.745 (0.019)** | <1e-99 | **-0.084 (0.027)** | 0.002 |
| **SDMT score<sup>a,c</sup>** | **0.217 (0.026)** | <1e-15 | **-0.488 (0.133)** | 2e-4 |
| **SWRT score<sup>a,c</sup>** | **0.390 (0.023)** | <1e-63 | **-0.368 (0.098)** | 2e-4 |
| **BMI<sup>a</sup>** | -0.025 (0.028) | 0.369 | **-0.183 (0.072)** | 0.011 |

<sup>a)</sup> All outcome measures and continuous predictor variables were standardized to a mean of zero and a standard deviation of one before inclusion in the models.
<sup>b)</sup> All effect estimates were obtained using the within-subject variance estimator by robust regression (WiSER) Julia package with default settings and 100 iterations (except for BMI, where 4 iterations were used, because more iterations resulted in non-convergence of the model).<sup>22</sup> For each of the six clinical outcome measures a separate model was run including sex, age, age<sup>2</sup>, mutant CAG repeat size and its interaction with age, and residual age at onset as fixed-effects, an intercept and slope for age as random effects, and sex, age, mutant CAG repeat size and its interaction with age, and residual age-at-onset as within-subject variability predictors. Effect estimates represent standardized regression coefficients (standard error) with all statistically significant effect estimates highlighted in bold.
<sup>c)</sup> Both Unified Huntington Disease Rating Scale cognitive scores indicate the total number of correct items (in 1 minute).
**Abbreviations:** BMI = body mass index, CAG = cytosine-adenine-guanine, SD = standard deviation, SDMT = symbol digit modalities test, SWRT = Stroop word reading test, WS = within-subject

**Figure 2.**
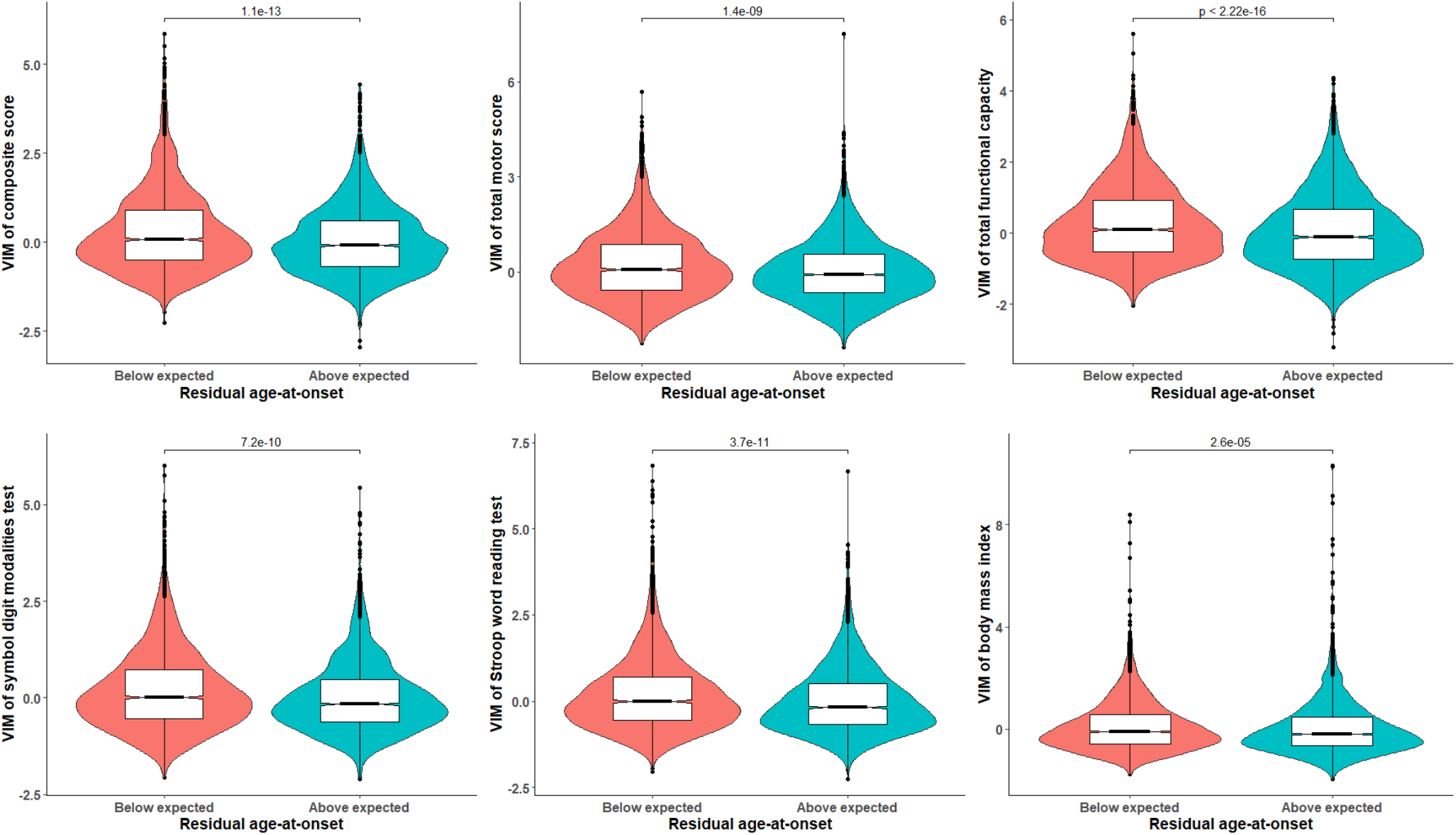
Younger age-at-onset is associated with increased within-individual clinical variability in Huntington disease. The violin plots depict the association of residual age-at-onset (i.e., whether the observed age-at-onset was below or above expected age of onset based on mutant *HTT* CAG repeat size) and the standardized variance independent of the mean (VIM) metric. The boxes indicate the interquartile ranges around the median (thick black lines), with values deviating more than 1.5 times the interquartile range from the median represented by black dots. After regressing out the effects of sex, age at baseline, mutant CAG repeat size and its interaction with age at baseline, and domain-specific disease severity at baseline from the VIM scores, the residuals were compared across the two categories of residual age-at-onset using the non-parametric Wilcoxon’s rank sum test.

### Part 2: Within-individual variability within the Bayesian brain theory framework

Increased variability in phenotype across time in a given individual may reflect the progressive breakdown of homeostatic mechanisms, and thus increased entropy, due to accruing pathology at molecular, cellular, tissue, organ or system level. To assess the validity of this hypothesis, the ‘free energy principle’-based Bayesian brain model, which is thought to provide a unified theory of brain function,^24^ was leveraged. Within this framework, phenotype was defined here as the “action” of the agent, because this is the only perceptible part of the variational free energy minimization process that could be measured by an external observer (e.g., a clinical rater).

#### Analytical derivation of the relation between entropy and phenotypic variability

Specifically, it was assessed how action is related to changes in the entropy of the underlying distribution, which for Gaussian distributions is directly related to the variance *σ* (Note: for consistency, the same terminology and symbols as used by Buckley et al.^25^ are employed, and for clarity of presentation, the derivation is restricted to the univariate case). Let *x* be a random variable with a Gaussian distribution:

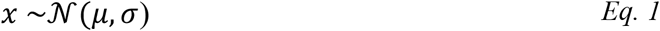

Then the differential entropy (*H*) of *x* is given by^26^:

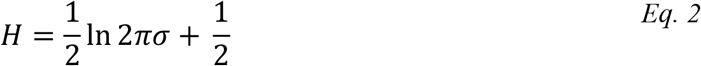

For a simple agent-based model possessing only a single brain state variable (*μ*) and a single sensory channel (*φ*), the organism believes its sensory input is generated by some generative function *g* parametrized by *θ*:

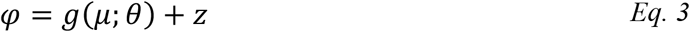

With *z* denoting random noise with zero mean and variance σ_*z*_, and *μ* denoting the organism’s beliefs about how the environmental states are generated, as follows:

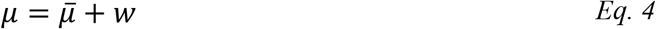

Here 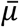 represents some fixed parameter, while *w* denotes random noise with zero mean and variance *σ*_*w*_. Assuming that with accruing neuropathology the estimates of the organism’s beliefs regarding its environment become increasingly unstable and unpredictable, which can be taken to imply that the variance of the distribution from which *w* arises increases, in the following I will thus specifically seek to derive the explicit function describing the relation between action (*a*) and *σ*_*w*_. For a simple agent-based model, *a* is given by^25^:

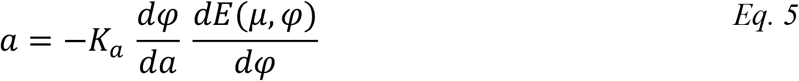

From which it follows that:

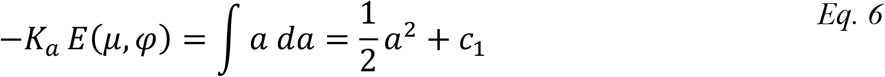

Here *K*_*a*_ denotes the learning rate associated with the action (with the negative sign indicting that the direction of action is such to oppose departures from steady-state), and *E*(*μ, φ*) represents the Laplace-encoded energy given by^25^:

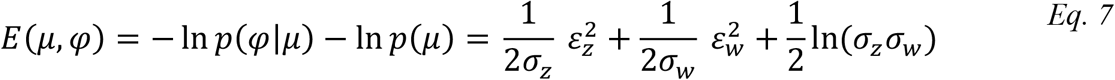

Where:

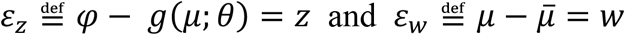

Therefore, to assess how action changes as a function of entropy, one needs to take the partial derivative of *Eq. 7* with respect to *σ*_*w*_:

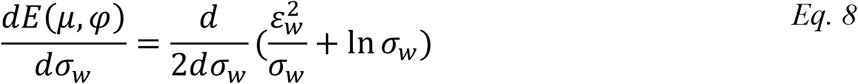

Now, let:

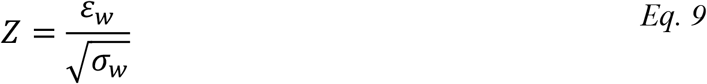

Substituting *Eq. 9* in *Eq. 8* yields:

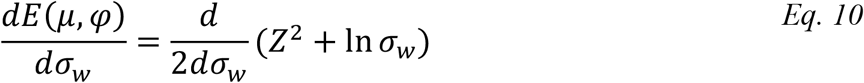

Given that *Z*, by definition, is scaled to the standard normal distribution, i.e., *Z* ∼ 𝒩(0, 1), and thus independent of the choice of *σ*_*w*_, *Eq. 10* simplifies to:

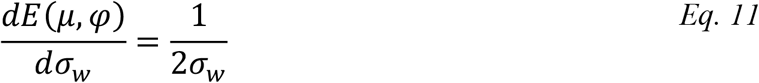

From which it follows that:

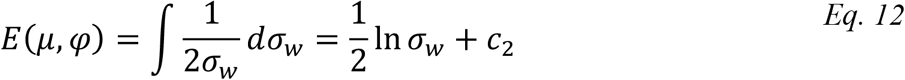

Combining *Eq. 6 & Eq. 12* (and setting the integration constants *c*_1_ = *c*_2_ = 0) obtains:

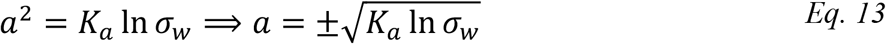

*Eq. 13* thus indicates that the *magnitude* of the action towards the steady-state is a monotonously increasing function of *σ*_*w*_ (with its direction opposite to that of the deviation from the steady-state), and by implication (given *Eq. 2*) also a monotonously increasing function of the entropy of the underlying distribution from which *w* (or *ε*_*w*_) arises. As 𝔼(*a*) = 0 during steady-state (because, by definition (*Eq. 4*), during a steady-state any deviation from the set-point 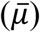 will be due to random noise *w*, which the action *a* tends to oppose), the variance of the action *a* can simply be derived as:

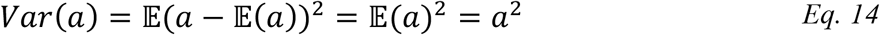

From *Eq. 13* it then follows that: *Eq. 14*

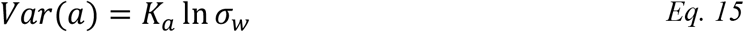

*Eq. 15* Therefore, *Eq. 15* demonstrates that variability of action, a measure of within-individual variability, is a monotonously increasing function of *σ*_*w*_, and thus of the entropy of the encoded brain states. This implies that within-individual variability in phenotype could be regarded as an increase in the entropy of the output of the underlying neuronal substrates encoding the organism’s beliefs about its environment.

#### Simulations using a Bayesian agent support link between entropy and phenotypic variability

To illustrate the analytically derived relations above, the previously published code for simulating a simple but complete agent-based model that utilizes the free-energy principle to perform perceptual inference^25^ was leveraged (the R code of the implementation is provided in the **Supplementary Materials**). Briefly, this agent’s environment consists of a unidimensional line and a single temperature source. The agent’s position on this line is represented by the environmental variable *ϑ* and its temperature *T* depends on its position as follows:

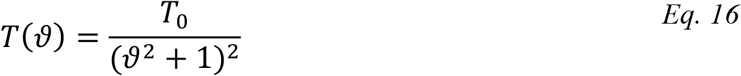

This agent is presumed to sit on a frictionless line and be stationary in the absence of action. The agent is allowed to set its own velocity, which is defined as its action *a*:

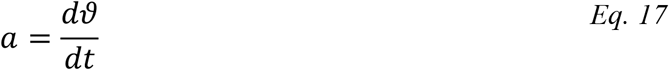

Further details are provided in the original publication.^25^ Here the behavior of this agent was simulated to assess whether an increase of the entropy of the distribution that underlies the agent’s beliefs about its environment (parametrized through *σ*_*w*_), would lead to an increase in the variability of the agent’s movements (represented by *Var*(*a*)), as predicted by *Eq. 15*. To this end, the agent’s desired and initial temperature were both to set to 20 °C, and its action for a time period of 1000 arbitrary units was simulated for different values of *σ*_*w*_, ranging from 1 to 3 (the exact initialization parameters/values and further details to reproduce all simulation results can be found in the R script provided in the **Supplementary Materials**). The magnitude of action was scaled to the variance of the sensory input *σ*_*z*_. **Figure 3** illustrates the agent’s action as a function of time for three different values of *σ*_*w*_, indicating increased variability of action for higher values of *σ*_*w*_. This latter association was closely in line with the analytical predictions and could be directly confirmed by calculating the non-parametric Spearman’s correlation coefficients between *σ*_*w*_ and estimates of the variability of the simulated actions (Spearman’s *ρ* = 0.95, p < 2.2e-16; **Supplementary Figure 3** and **Figure 4** (top panel), all correlations were calculated for the steady-state defined as time ≥ 750). Given that, in this particular case, the agent’s actions wholly consisted of ‘measurable’ movements, it was also possible to readily calculate the work performed by the agent, defined as the cumulative sum of the product of acceleration and distance traveled over time. Importantly, this analysis demonstrated that the amount of work required to maintain steady-state is strongly associated with *σ*_*w*_ (Spearman’s *ρ* = 0.97, p < 2.2e-16; **Figure 4**, bottom panel). Thus, these simulations confirmed the analytically derived relations presented in the previous section, illustrating that increased entropy of brain states not only will lead to higher phenotypic variability, but will also increase the energy costs required to maintain steady-states.

**Figure 3.**
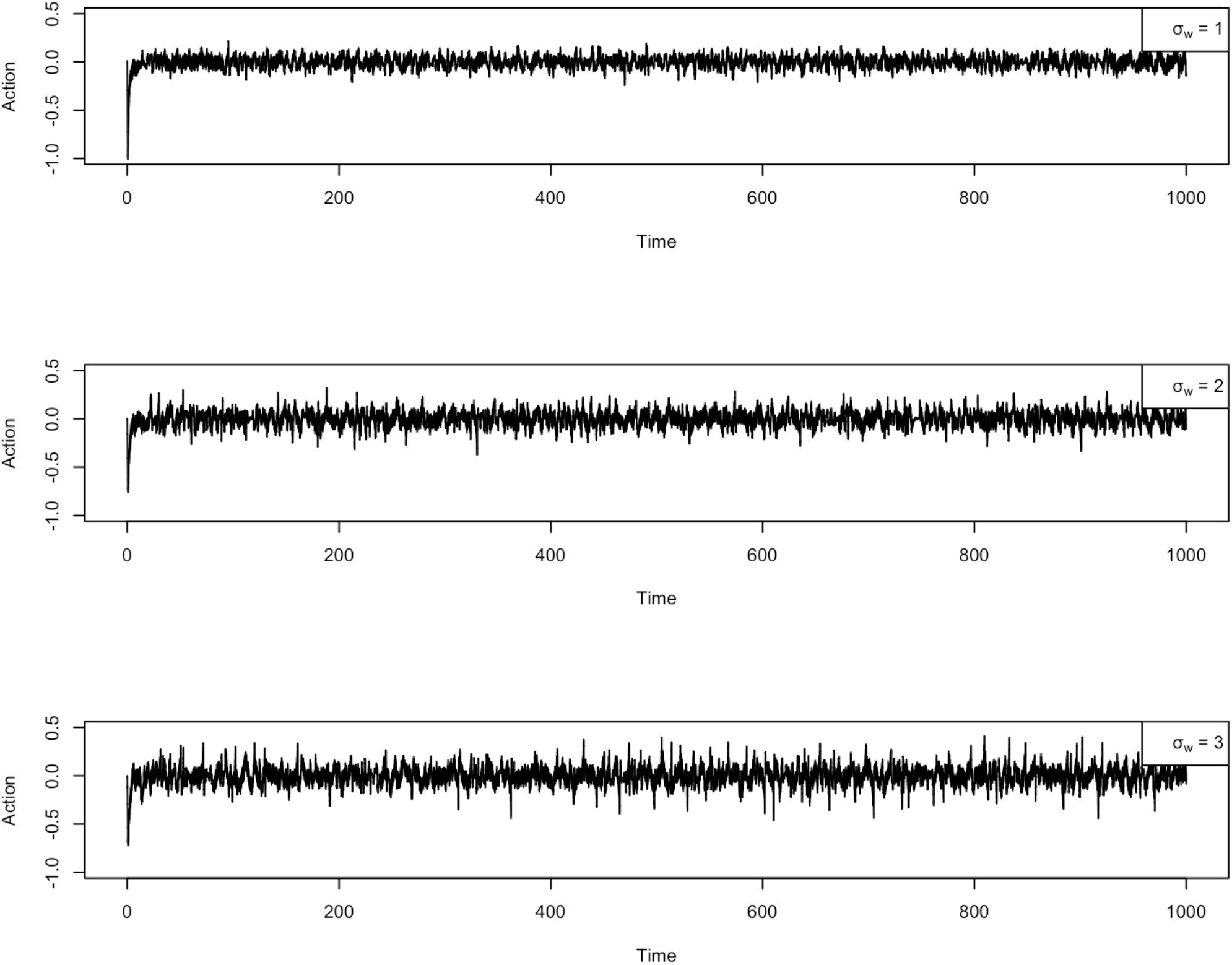
Phenotypic variability increases with higher values of predictive entropy. The figure depicts the simulated action (i.e., movements around a predefined set-point) of a simple Bayesian agent for different values of the entropy of the distribution that underlies the agent’s beliefs about its environment as parametrized through *σ*_*w*_. Variability of action is larger for higher values of *σ*_*w*_. See main text for more details.

**Figure 4.**
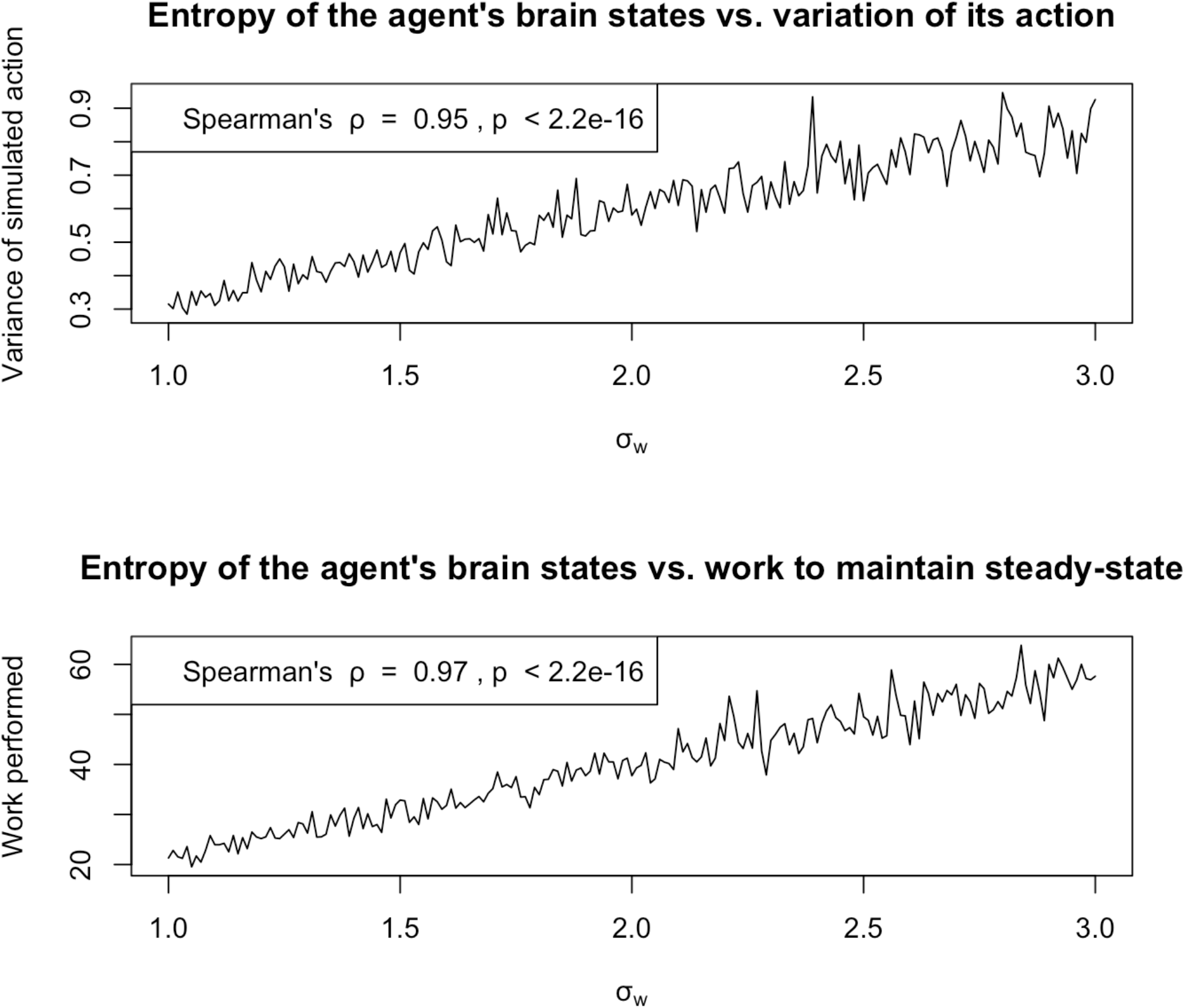
Increased phenotypic variability results in higher energy expenditure. The agent’s phenotypic variability increased with higher values of the entropy of the distribution that underlies the agent’s beliefs about its environment as parametrized through *σ*_*w*_ (top panel). The amount of work performed by the agent required to maintain steady-state – defined as the cumulative sum of the product of acceleration and distance traveled over time – increased with higher values of *σ*_*w*_ (bottom panel).

### Part 3: Leveraging variability to gain insights into Huntington disease pathogenesis

This section seeks to demonstrate the utility of accounting for phenotypic variability as an integral part of HD. Specifically, it is illustrated how considering variability as a core feature of HD could provide new insights into the pathogenesis of weight loss, as well as facilitate the uncovering of novel genetic modifiers of the disease.

#### Weight loss

Weight loss is a hallmark of HD, is seen in both patients and most genetic (rodent) models of the disease, can precede the onset of motor signs and symptoms by many years, and is a robust predictor of the rate of disease progression.^27, 28^ However, its precise underlying causes are poorly understood.^29^ As detailed in the previous section, increased within-individual variability in phenotype is inevitably related to higher energy expenditure, because more work will be required to maintain steady-states in the face of higher entropy of the brain’s predictive machinery as a result of pathology. Applied to HD, this theoretical framework predicts that increased phenotypic variability would result in increased energy expenditure, and thus a lower body weight and a higher rate of weight loss. To directly probe this predicted association, the within-individual variability in the UHDRS total motor score was assessed in relation to both body weight and the rate of weight loss in the Enroll-HD cohort. Average body weight (estimated through a random intercept linear mixed-effects model with adjustment for age, age^2^, sex, mutant CAG repeat size, as well as its interaction with age, *and total motor score*) decreased with higher within-individual variability of total motor score (**Figure 5**, left panel). Moreover, the rate of body weight loss (represented by the random slope coefficients obtained from a random intercept and slope linear mixed-effects model with age, age^2^, sex, mutant CAG repeat size, as well as its interaction with age, *and total motor score*, as fixed effects, and an intercept and age as random effects) also increased with higher variability of total motor score (**Figure 5**, right panel). These findings thus indicate that higher energy expenditure in HD may not be necessarily due to an inherent defect in systemic energy regulation, but could be an inevitable consequence of increased energy requirements to maintain homeostasis in the face of a progressively inefficient capacity for predictive inference.

**Figure 5.**
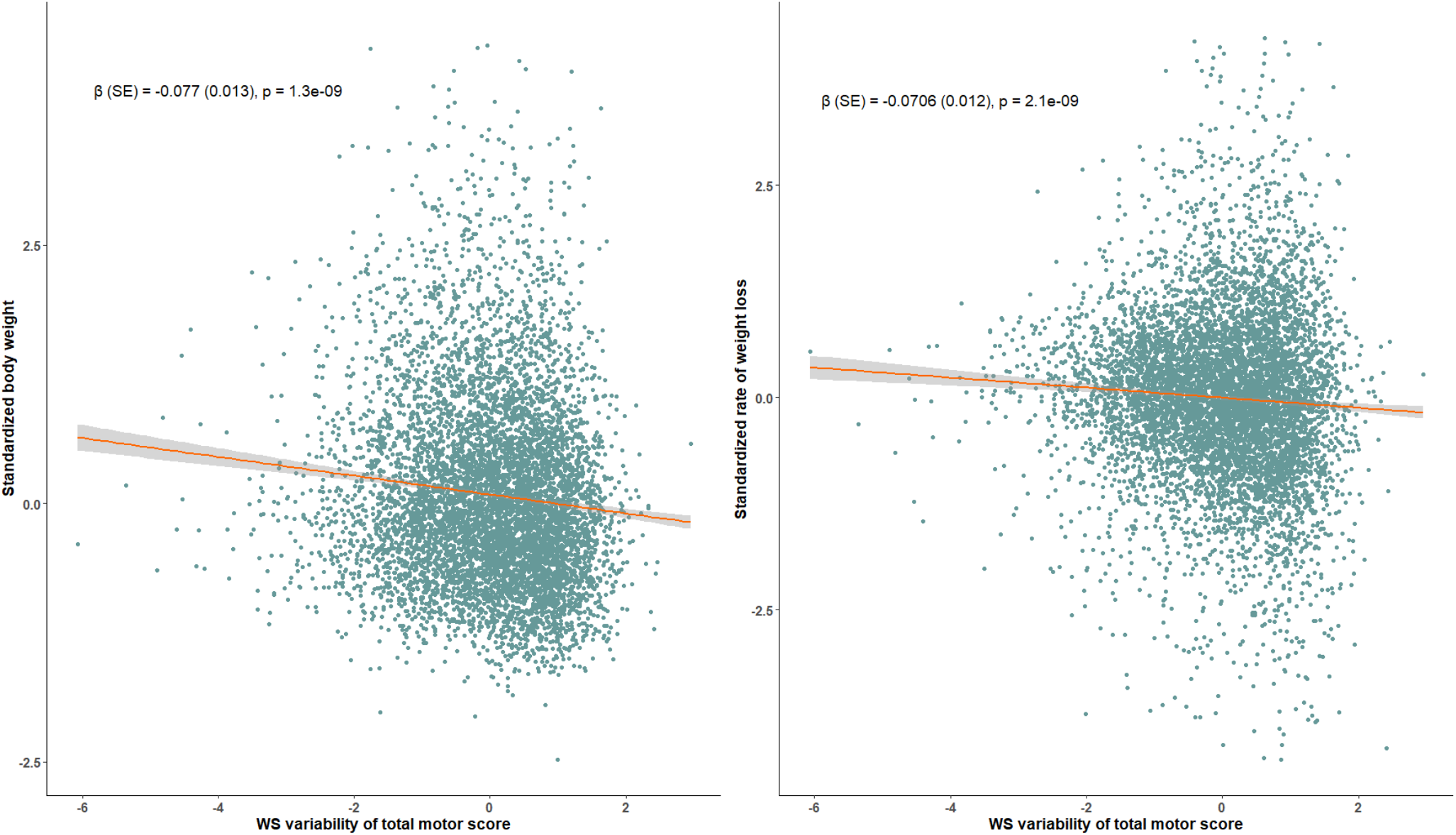
Within-individual variability of motor impairment is associated with lower body weight and a faster rate of weight loss. Average body weight (estimated through a random intercept linear mixed-effects model with adjustment for age, age^2^, sex, mutant CAG repeat size, as well as its interaction with age, *and total motor score*) decreased with higher within-individual variability of total motor score (left panel). Similarly, the rate of body weight loss (represented by the random slope coefficients obtained from a random intercept and slope linear mixed-effects model with age, age^2^, sex, mutant CAG repeat size, as well as its interaction with age, *and total motor score*, as fixed effects, and an intercept and age as random effects) also increased with higher variability of total motor score (right panel). The insets indicate the standardized regression estimates (standard error (SE)). The red lines represent the regression lines, with the corresponding grey areas indicating the SEs around the mean. WS = within-subject.

#### Genetic modifiers of within-individual variability

The GWAS of within-individual variability of BMI identified a genome-wide significant intergenic locus on chromosome 22, tagged by the lead SNP rs62231080 (standardized β = - 0.260 ± 4.57E-2, p = 1.48E-8) (**Table 3, Figure 6**, and **Supplementary Figure 3**). Positional and eQTL analyses mapped this locus to the protein coding genes *NUP50, FAM18A*, and *RIBC2*, and the lincRNA genes *CTA-217C2*.*1* and *CTA-268H5*.*14* (**Figure 6**). The GWAS of mean BMI levels revealed two additionally novel loci on chromosomes 16 and 18, tagged by the lead SNPs rs200717776 and rs11874212, respectively (**Table 3, Figure 7**, and **Supplementary Figure 4**). Positional and eQTL analyses mapped the chromosome 16 locus to the protein coding *HEATR3* gene, and the antisense lncRNA *RP11-429P3*.*5*, and the chromosome 18 locus to the protein coding *TSHZ1* and *RP11-17M16*.*1* genes (**Figure 7**). Notably, two independently significant SNPs (rs11150911 and rs12964145) at the chromosome 18 locus have previously also been associated with BMI and body weight in population-based cohorts.^30, 31^ Moreover, MAGMA gene-based analysis identified *EIF3F* as a gene associated with mean BMI levels in HD patients (gene-based p-value = 2.03E-6 (< Bonferroni-corrected threshold of 0.05/19208 = 2.60E-6), while another gene on chromosome 14 (*KLHL33*) showed only suggestive statistical significance (p = 3.48E-6) (**Supplementary Figure 5**). Tissue and pathway enrichment analysis did not yield any additional significant associations (data not shown).

**Table 3.** Genetic loci associated with within-individual variability or mean levels of body mass index, total motor score or symbol digit modalities test scores in Huntington disease.

| Trait | Outcome | Chr | Lead SNP | BP (hg19) | Effect allele | MAF (%) | Effect size<sup>a</sup> | SE | P-value | Candidate modifier genes |
| --- | --- | --- | --- | --- | --- | --- | --- | --- | --- | --- |
| **BMI** | *variability* | **22** | **rs62231080** | **45484152** | **A** | **21.8** | **-0.260** | **4.57E-02** | **1.48E-08** | ***NUP50*** |
|  | *mean* | **16** | **rs200717776** | **50145334** | **CT** | **10.1** | **0.408** | **7.35E-02** | **3.43E-08** | ***HEATR3*** |
|  |  | **18** | **rs11874212** | **73503347** | **A** | **30.1** | **0.222** | **3.90E-02** | **1.59E-08** | ***TSHZ1*** |
| **TMS** | *variability* | 16 | rs772972 | 27838619 | T | 32.4 | -0.188 | 3.68E-02 | 4.10E-07 | *GSG1L* |
|  | *mean* | 3 | rs1984782 | 153289784 | G | 46.3 | 0.125 | 2.50E-02 | 6.21E-07 | *P2RY1* |
|  |  | 6 | rs36079127 | 99300452 | G | 37.9 | -0.134 | 2.55E-02 | 1.93E-07 | *FBXL4* |
| **SDMT** | *variability* | 1 | rs4101233 | 91238773 | T | 38.5 | 0.204 | 3.85E-02 | 1.26E-07 | *CDC7, ZNF644* |
|  | *mean* | **5** | **rs34322595** | **79998532** | **T** | **31.2** | **-0.151** | **2.64E-02** | **1.30E-08** | ***MSH3*** |
|  |  | 12 | rs55833733 | 125839198 | T | 22.2 | 0.146 | 2.74E-02 | 1.16E-07 | *TMEM132B* |
|  |  | 13 | rs9559664 | 110471106 | G | 15.3 | -0.175 | 3.41E-02 | 3.45E-07 | *IRS2, RN7SKP10* |
|  |  | 15 | rs55666775 | 100431982 | A | 9.5 | -0.217 | 4.07E-02 | 1.09E-07 | *CTD-2054N24.2, CTD-3076O17.1* |

<sup>a</sup>) Effect estimates represent standardized regression coefficients. All loci that passed the genome-wide (p-value < 5E-8) or suggestive (p-value < 1E-06) statistical significance threshold are summarized. Genome-wide significant loci are marked in bold.
**Abbreviations:** Chr = chromosome, BP = base pair position in Human Genome version 19 coordinates, BMI = body mass index, MAF = minor allele frequency, SDMT = symbol digit modalities test, SE = standard error, TMS = total motor score.

**Figure 6.**
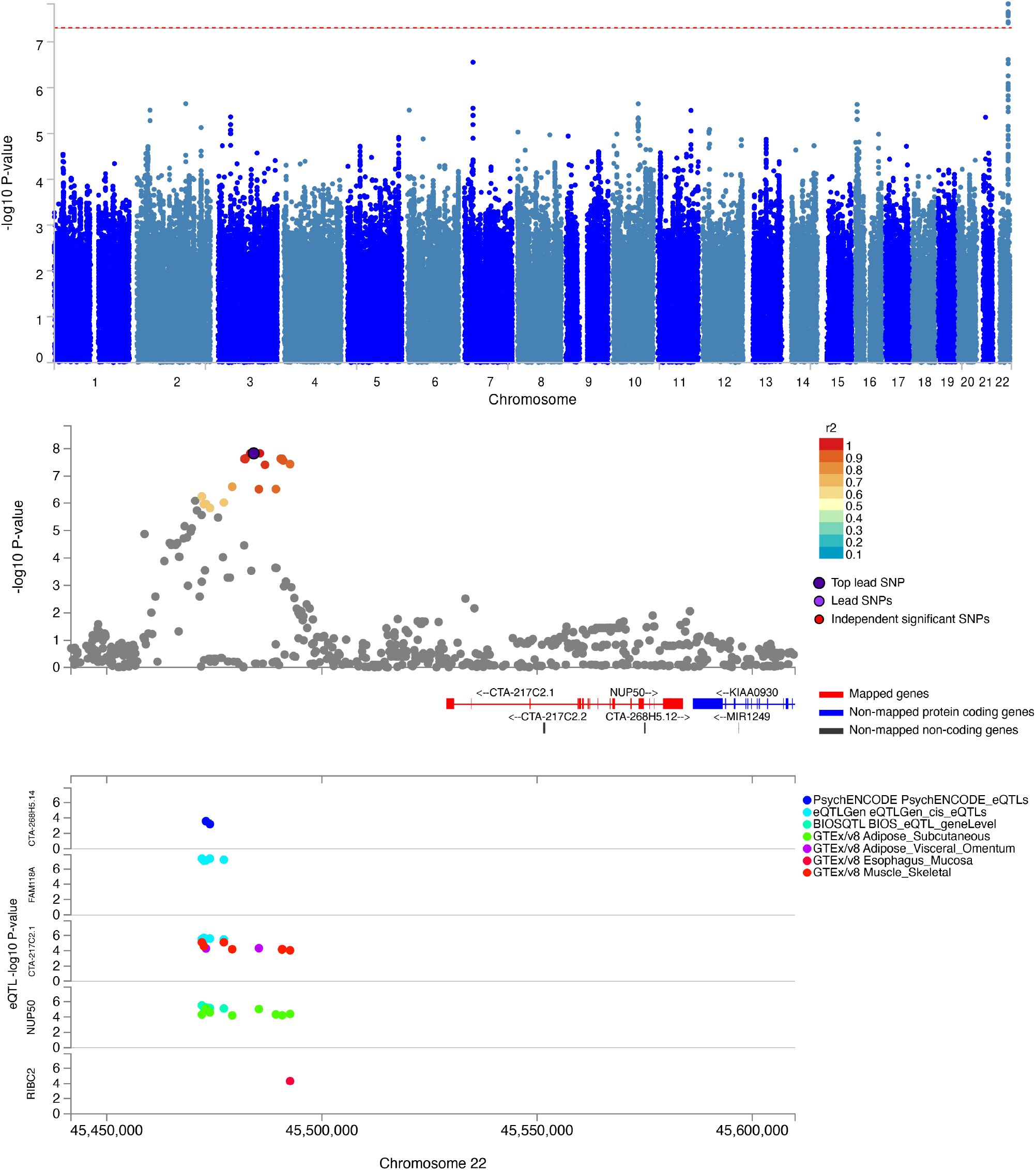
Genome-wide association study of body mass index variability in Huntington disease. The dashed red line in the Manhattan plot (top plot) indicates the threshold for genome-wide significance (i.e., p = 5.0E-8). An intergenic locus on chromosome 22, tagged by the lead SNP rs62231080, reached genome-wide significance (standardized β = -0.260 ± 4.57E-2, p = 1.48E-8). Positional (middle panel) and eQTL (bottom panel) analyses mapped this locus to the genes *NUP50, FAM18A*, and *RIBC2*, and the lincRNA gene *CTA-217C2*.*1* and *CTA-268H5*.*14*.

**Figure 7.**
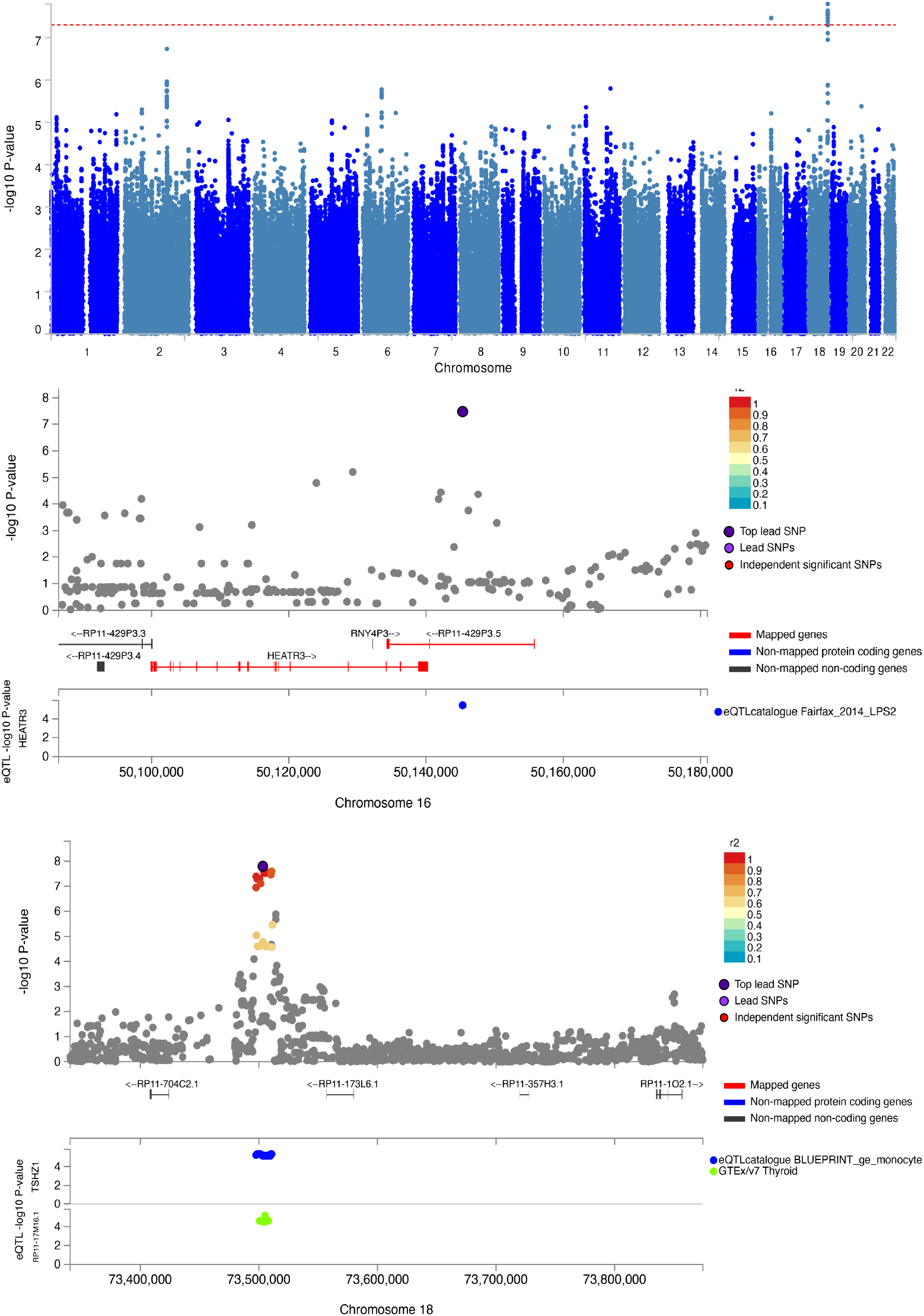
Genome-wide association study of mean body mass index in Huntington disease. The dashed red line in the Manhattan plot (top plot) indicates the threshold for genome-wide significance (i.e., p = 5.0E-8). Two loci on chromosomes 16 and 18, tagged by the lead SNPs rs200717776 and rs11874212, respectively, reached genome-wide significance. Positional and eQTL analyses (lower panels) mapped the chromosome 16 locus to the protein coding *HEATR3* gene, and the antisense lncRNA *RP11-429P3*.*5*, and the chromosome 18 locus to the protein coding *TSHZ1* and *RP11-17M16*.*1* genes.

The GWAS of within-individual variability of UHDRS total motor score revealed a locus on chromosome 16 (rs772972) at suggestive significance (p = 4.10E-7), which was located in an intronic region of the *GSG1L* gene (**Supplementary Figure 6**). Interestingly, this locus has previously been associated with residual age-at-onset in HD.^16^ The GWAS of mean UHDRS total motor score did not identify genome-wide significant loci (**Table 3**). The GWAS of within-individual variability of UHDRS SDMT revealed a locus on chromosome 1 (rs4101233) at suggestive significance (p = 1.26E-7), which was located in an intergenic region (**Supplementary Figure 7**). Notably, this locus could be mapped to the protein coding genes *ZNF644* and *CDC7*, the latter of which encodes a DNA replication enzyme that may be related to *FAN1* activity.^32^ Only the well-characterized *MSH3* locus on chromosome 5, which has previously been associated with residual age-at-onset in HD,^16^ reached genome-wide significance in the GWAS of mean SDMT levels (**Table 3**).

#### Genetic modifiers of age of onset

Given that age of onset in HD is usually recorded as a single measurement, it is not possible to directly assess within-individual variability of age of onset. However, in Part I of the results section, it was shown that within-individual variability is strongly associated with mutant CAG repeat size. Given that within-individual variability is also likely to account for at least part of the between-individual variability, here the association between CAG repeat size and between-individual variability in age of onset is examined in closer detail, using the same dataset consisting of 9064 individuals compiled by the GemHD consortium.^16^ At first glance, the between-individual variability of residual age-at-onset appears to *decrease* for larger CAG repeat sizes (**Figure 8**, left panels; Spearman’s *ρ* = -0.91, p < 0.001). However, given the strong inverse association between mutant CAG repeat size and age of onset, this association is likely to be confounded by the substantially younger age of onset of the participants with larger CAG repeat sizes. To account for this confounding by age, residual age-at-onset was divided by the predicted age of onset (based on mutant CAG repeat size). Indeed, after accounting for predicted age of onset, the variability of the *relative* residual age-at-onset (i.e., expressed as a fraction of the predicted age of onset) strongly *increased* with larger mutant CAG repeat size (**Figure 8**, right panels; Spearman’s *ρ* = 0.81, p < 0.001).

**Figure 8.**
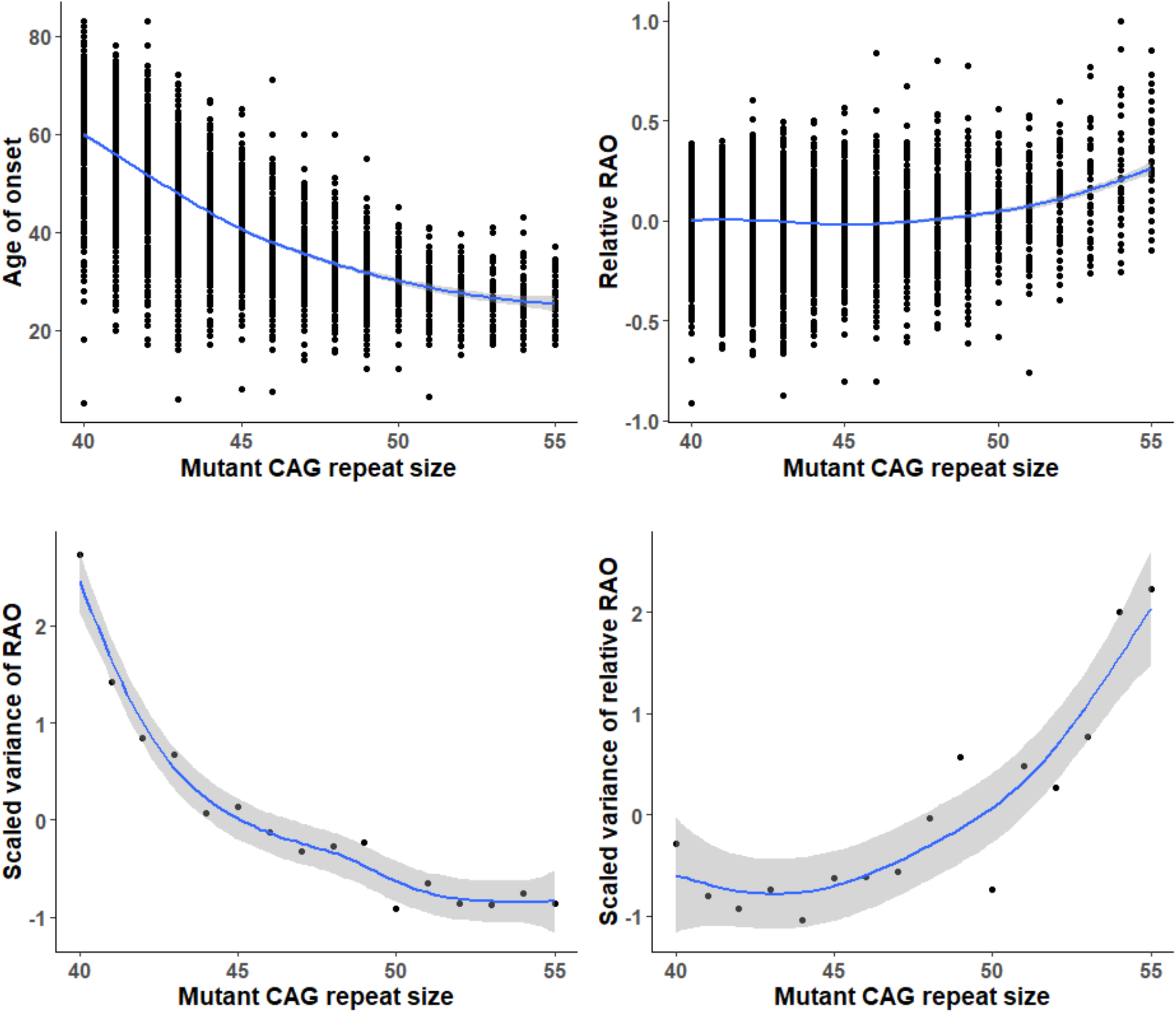
Age of onset variability in Huntington disease. Although both absolute age of onset and residual age-at-onset (RAO) variability *decrease* with higher mutant CAG repeat size (left panels), the relative RAO, as well as its variability *increase* with larger mutation sizes (right panels). The blue lines represent smoothing splines and associated standard errors around the mean indicated in grey. See main text for more details.

Due to the strong association between CAG repeat size and the rate of disease progression (or age of onset as a proxy for the rate of disease progression),^3^ it is obvious that delaying or hastening of age of onset by a certain number of years cannot be assumed to indicate the same degree of disease modification for individuals with different mutant CAG repeat sizes (see **Appendix** for an explicit analytical explanation). For example, assuming that pathological burden increases linearly from birth onwards, to delay the expected age of onset by 10 years in two premanifest mutation carriers with expected ages of onset of 30 and 40 years, the rate of disease progression would need to be lowered by 25% and 20%, respectively (**Appendix**). Therefore, using the relative difference between the actual and expected age of onset (as opposed to the absolute difference between actual and expected age of onset as utilized previously^16^), is likely to result in more accurate estimates of potential genetic modifiers of age of onset in HD.

To directly assess this postulate, a GWAS of the *relative* residual age-at-onset in HD was conducted, using the same dataset and otherwise exact same covariates and approach as previously published by the GemHD consortium.^16^ Indeed, this approach resulted in the identification of a substantially larger number of genome-wide significant SNPs than reported previously (i.e., 488 *vs*. 328 (excluding those on chromosome 4 as these may reflect artefacts due to rare CAG repeat sequence haplotypes^16^)) (**Figure 9**). Interestingly, almost all (∼98%) genome-wide significant SNPs identified previously were also detected using relative residual age-at-onset as the outcome (**Supplementary Figure 8**), suggesting a sizable increase in statistical power.

**Figure 9.**
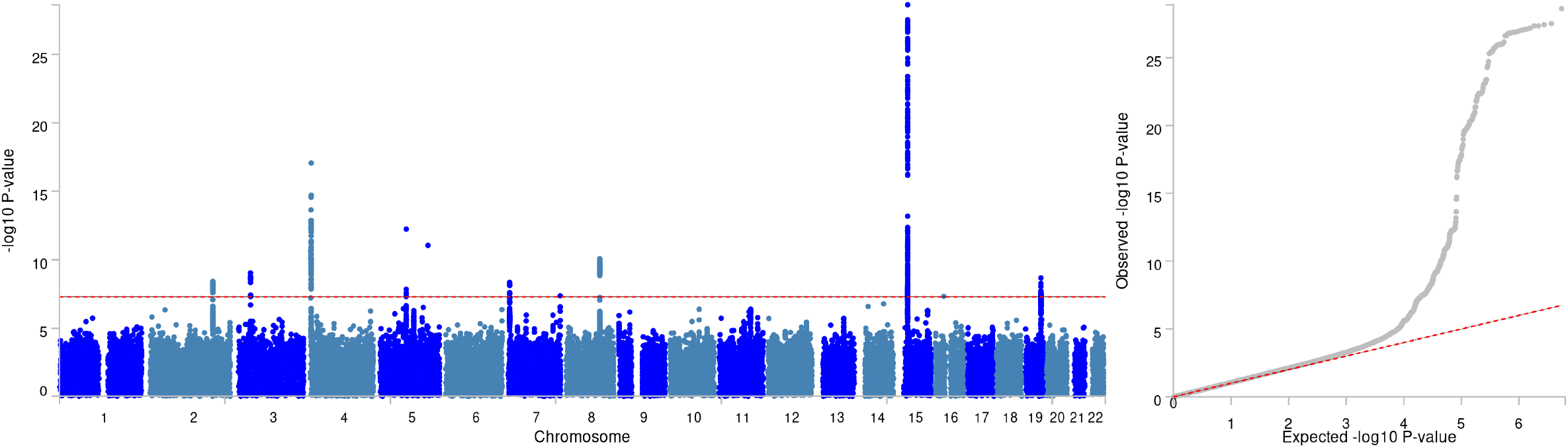
Genome-wide association study of *relative* residual age-at-onset in Huntington disease. The Manhattan (left) and corresponding quantile-quantile plot (right) are depicted. The dashed red line in the Manhattan plot indicates the threshold for genome-wide significance (i.e., p = 5E-8). See main text and **Table 4** for more details about the identified loci.

To allow direct comparisons, for all potential genetic modifiers reported previously,^16^ the effect estimates and associated p-values were compared between the two methods (**Table 4**). Although all effect estimates were directionally consistent and mostly similar between the two methods, using relative residual age-at-onset resulted in the identification of an additional novel genome-wide significant locus tagged by a rare variant (rs138433183) on chromosome 7, located in an intronic region of the *ESYT2* gene (**Figure 10**). Moreover, three other loci that failed to reach genome-wide significance before using residual age-at-onset as the outcome (including the *PMS1* and *PMS2* loci, as well as another locus on chromosome 19 (19AM3)), reached genome-wide significance using relative residual age-at-onset as the outcome (**Table 4**). Conversely, the *SYT9* locus on chromosome 11 did not reach genome-wide significance using the current method, suggesting a potential false positive hit in the previous GWAS. More in-depth characterization of these loci is beyond the scope of this paper and can be accomplished by interested researchers using the provided FUMA output files and summary statistics (**Supplementary Materials**).

**Table 4.**
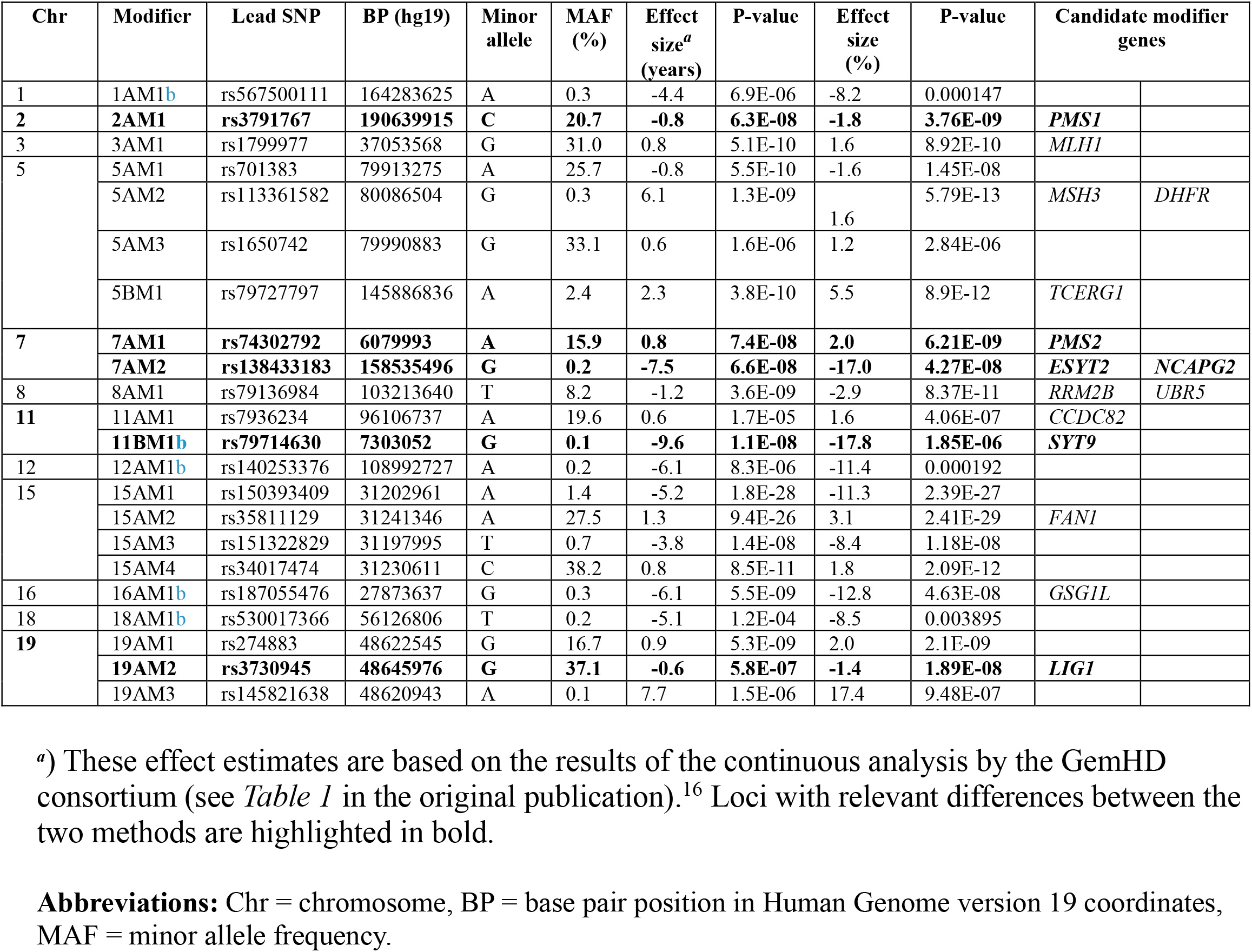
Comparison between absolute vs. relative residual age-at-onset genetic modifiers.

**Figure 10.**
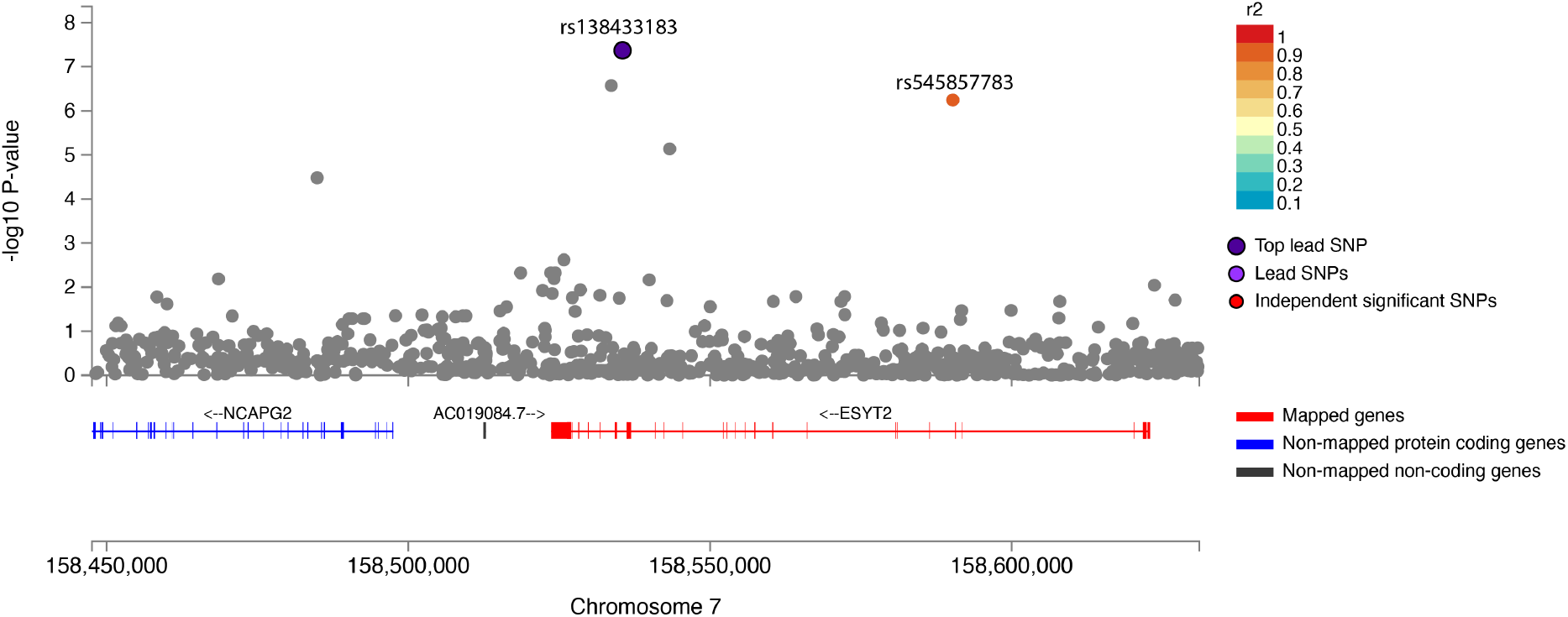
Genome-wide association study of *relative* residual age-at-onset in Huntington disease identifies a novel locus. The figure depicts the genomic location of novel rare variants on chromosome 7 within the *ESYT2* gene, tagged by the lead (rs138433183) and an independently significant (rs545857783) SNP, associated with *residual* age-at-onset in Huntington disease.

## Discussion

This is the first study to assess visit-to-visit within-individual variability and its clinical and genetic correlates in HD. Leveraging detailed genetic and longitudinal clinical data from large cohorts of HD patients, this work *1)* establishes within-individual variability in disease expression as an integral part of HD semiology, demonstrating that it increases across all clinical domains with disease duration, mutation size, younger age-at-onset, and lower body weight, *2)* provides a mathematical framework linking higher phenotypic variability to increased predictive entropy, revealing a fundamental relation between within-individual variability and energy expenditure, and *3)* identifies novel genetic modifiers of both within-individual variability and age-at-onset in HD, detailing new concepts and approaches that could also increase the statistical power of future genetic association studies in HD.

Despite the clinical impression that HD patients exhibit considerable variability in symptoms and signs independent of the measuring method or rater, no previous studies have targeted this particular aspect of HD. Here it was found that all clinical domains of HD, including motor, functional, cognitive and metabolic features of the disease, exhibit pronounced visit-to-visit variability. Moreover, within-individual variability robustly increased with larger mutant CAG repeat size for the composite, motor and functional scores, as well as BMI. For cognitive scores, within-individual variability tended to increase for CAG repeat sizes in the lower pathogenic range, followed by a leveling off of the effect for larger CAG repeat sizes. Given the strong association between mutation size and cognitive deterioration,^3^ this latter finding may thus be due to floor/ceiling effects of the cognitive testing batteries in HD.^23^ In addition, an age of onset lower than expected based on mutation size was also robustly associated with a higher degree of within-individual variability across all clinical domains of HD. Collectively, these findings thus establish within-individual variability in disease expression as an integral part of HD semiology.

Interestingly, motor impersistence, referring to the inability to sustain voluntary muscle contractions, has long been recognized as a cardinal feature of HD, and is thought to underlie other well-known clinical signs of the disease, including the ‘milkmaid’s sign’ (rhythmic squeezing of the examiner’s fingers), and the ‘fly-catcher’s tongue’ (inability to maintain tongue protrusion beyond lips).^33^ Albeit the precise cause of motor impersistence in HD is not fully elucidated, it could also be regarded as a manifestation of increased within-individual variability, though on a much smaller time-scale. Indeed, higher quantitative motor measures of variability – including grip and tongue force variability, as well as variability in inter-onset intervals during speeded tapping – have been reported in HD mutations carriers.^34^ In line with the findings of the current paper, increased variability on these quantitative motor measures was shown to be predictive of a faster rate of progression in early HD, while higher variability in inter-onset intervals during speeded tapping was also associated with a higher risk of disease onset in premanifest HD mutation carriers.^34^

Leveraging the free energy principle, which conceptualizes action as a means through which the brain attempts to minimize sensory prediction errors (i.e., ‘active inference’),^24^ the current work also provides a useful theoretical framework for understanding the potential causes of phenotypic variability in HD. Specifically, both analytically and through simulation experiments, it demonstrates that an increase in predictive entropy of the brain’s machinery (e.g., due to gradual breakdown of homeostatic mechanisms as a consequence of progressive neuronal dysfunction and loss) will inevitably lead to increased variability in phenotype. Although an exhaustive discussion of the free energy formulation is beyond the scope of this paper, here it is important to stress that within this framework ‘action’ is not necessarily limited to macroscopic, observable movements, but could encompass any active motion, including, e.g., secretory activity of internal (hormonal) glands, gastrointestinal motility, as well as modulation of the oculomotor and cardiorespiratory system. Interestingly, previously we found a higher ‘approximate entropy’ of prolactin secretion in early-stage HD patients compared to age, sex and BMI matched healthy control subjects,^35^ further supporting the notion of within-individual variability as a general feature of the disease. It should be noted though that the proposed Bayesian framework for linking entropy of brain states to (endo)phenotypic variability will only hold under the assumption that the agent will have retained the capacity to engage in active inference. For example, once patients have become bedridden in advanced stages of the disease, motor variability will be low, not because of reduced predictive entropy, but simply due to a dramatic loss of motor control.

One common but still little understood feature of HD is progressive weight loss, which is strongly associated with mutation size, already starts in the premanifest stages of the disease despite adequate dietary intake, and has been associated with higher resting energy expenditure.^36-38^ Although a higher BMI is robustly associated with a slower rate of disease progression in HD,^27^ in a previous Mendelian Randomization study we demonstrated that this association is not causal, but likely a *consequence* of a slower rate of disease progression.^29^ Consistent with the proposed framework linking a higher predictive entropy to increased phenotypic variability, and consequently to increased energy expenditure, in the present study a higher variability of motor features of HD was indeed associated with both lower body weight and a faster rate of weight loss, *independent* of the severity of motor impairment. These findings thus indicate that higher energy expenditure in HD may not necessarily be due to inherent defects in systemic energy regulation, but rather could be an inevitable consequence of increased energy requirements to maintain homeostasis in the face of a progressively inefficient capacity of a diseased brain for predictive inference. In fact, this could also account for the ubiquity of weight loss in other neurodegenerative diseases, in which emaciation generally accompanies a faster rate of disease progression.^39^ Therefore, in the context of HD and other brain disorders, BMI and weight loss could be regarded as proxies for the underlying rate of neuropathology.

This work also demonstrates that accounting for phenotypic variability can accelerate the discovery of novel genetic modifiers of HD. First, a GWAS of within-individual BMI variability identified a novel genome-wide significant locus (rs62231080) near the *NUP50* gene, which was also associated with the expression levels of this gene. Interestingly, variants in *NUP50*, encoding the nucleopore basket protein NUP50, were recently also associated with amyotrophic lateral sclerosis, with loss of NUP50 affecting splicing regulation, neuronal survival and motor function in multiple animal models.^40^ Moreover, disruption of nucleocytoplasmic transport is a consistent feature of HD models and was recently also linked to neurodevelopmental defects in cellular and Drosophila models of HD.^41-43^ Notably, the rs62231080 locus was distinct from three other novel loci associated with mean BMI levels. Interestingly, *EIF3F*, which was identified through gene-based analysis and encodes a subunit of the eukaryotic translation initiation factor eIF3, has also been linked to neurodevelopmental defects.^44^ Importantly, eIF3F favors repeat-associated non-ATG (RAN) translation, a key mechanism through which repeat expansions could induce neurodegeneration, with its knockdown reducing the levels of RAN proteins.^45^ Given that both BMI and its variability could reflect the rate of neuropathology as argued above, these loci thus do not necessarily need to be involved in systemic energy regulation per se, but may also exert their influence by affecting neurodegeneration in HD. Similarly, GWASs of within-individual variability in the UHDRS TMS and SDMT scores, identified other novel loci, although these latter loci did not reach genome-wide significance likely due to the relatively small sample size available for the longitudinal genetic association studies. Second, by accounting for the association between mutation size and age of onset variability through the derivation of a new metric – i.e., relative residual age-at-onset – it was demonstrated that also the statistical power of GWASs of age of onset in HD can be increased considerably. Indeed, utilizing relative instead of absolute residual age-at-onset resulted in the identification of 49% more genome-wide significant SNPs, as well as the discovery of an additional genome-wide significant locus in the *ESYT2* gene, encoding the extended synaptotagmin 2 protein that is important for calcium-dependent lipid binding and transport. Interestingly, gene fusion between *HTT* and *ESYT2* has previously been linked to breast cancer.^46^ Additionally, given that extended synaptotagmin 2 mediates endocytosis of activated fibroblast growth factor receptors,^47^ and fibroblast growth factor treatment has been shown to be neuroprotective in HD animal models,^48^ alterations in the growth factor signaling pathway might partly underlie these observed effects.

Although due to the strong association between mutant *HTT* CAG repeat size and the rate of disease progression, as well as availability of large and comprehensive datasets, HD provided an excellent model for deriving and testing the theoretical framework presented in this paper, there is no pertinent reason to assume that the association between increased predictive entropy and phenotypic variability would be *specific* to HD. Indeed, the proposed theory provides an intuitive framework for explaining a range of other little understood phenomena across different neurological diseases, including the association of increased visit-to-visit variability in body weight and blood pressure and a faster rate of disease progression in patients with Alzheimer and Parkinson disease.^13, 14^ weight loss preceding neurodegeneration by many years in other neurodegenerative diseases,^39^ the association between body weight variability and a higher risk of cognitive impairment and dementia,^11^ as well as the robust associations of many neuron-specific genes and BMI.^49^ Similarly, the concept of relative age-at-onset is likely to be useful in the study of other disorders in which there is a strong association between genotype and age of onset, especially other repeat expansion disorders exhibiting an inverse association between mutation size and age of onset. Finally, it should be noted that the concept of a ‘Bayesian agent’ is not necessarily limited to multicellular organisms and could be extended to other complex model systems capable of active inference, including cells and organoids. Therefore, the proposed link between predictive entropy and phenotypic variability could also be probed in model systems of HD and other (neurodegenerative) diseases where any dynamically regulated ‘phenotype’ could serve as a potential read-out. An exhaustive discussion of these other topics is beyond the remit of the current paper, but will be elaborated on in future work.

The current work also has several potential limitations. First, the *intraindividual* rate of disease progression was modeled linearly in time by using mixed effects models with random intercepts and slopes for age, because inclusion of higher order random terms for age led to model non-convergence, likely due to the relative sparsity of within-individual longitudinal measurements to enable stable estimates of higher order random effects. However, all models included a quadratic term for age as a fixed effect, which accounts for non-linear changes of the mean of the outcome measures over time. Moreover, an alternative method based on VIM, which uses a non-linear exponential model to estimate variability independent of the mean,^9^ yielded similar results.

Therefore, it is highly unlikely that increases of within-individual variability with age or mutant CAG repeat size could be accounted for by exponential increases in the rate of disease progression. Second, the visit-to-visit within-individual variability estimates were solely based on clinical outcome measures, including motor, functional and cognitive measures and BMI, obtained at about yearly or longer intervals. More objective, quantitative measures, such as the Q-Motor testing battery and other wearable motor and cognitive testing paradigms,^34, 50, 51^ applied at shorter intervals and more frequently, are thus likely to be more sensitive and yield more accurate estimates of within-individual variability profiles over time. Nevertheless, given that even the relatively crude clinical outcome measures utilized here could robustly capture at least part of the within-individual variability in phenotype, the current results are likely to represent underestimates of the true magnitude of within-individual variability in HD. Third, the available sample size for the GWAS of within-individual variability in HD was relatively small. Despite this, a genome-wide significant locus associated with within-individual variability of BMI was identified, while several other loci exhibited suggestive significance for within-individual variability of BMI, as well as motor and cognitive scores. Genotyping of a larger portion of the Enroll-HD cohort will be highly instrumental in identifying more genetic modifiers of within-individual variability in HD.

In conclusion, within-individual variability in disease expression is an integral part of HD semiology, accounting for which could facilitate the discovery of pathogenic mechanisms and outcomes directly relevant to the development of disease modifying therapies. Future studies should assess the utility of within-individual variability as an additional outcome measure: *1)* For the (re-)interpretation of the findings of past and current clinical trials, as well as for the design of future (clinical) studies, *2)* As a translatable outcome measure in cell and animal studies, as by definition it provides a scale invariant read-out sensitive to the earliest stages of pathology, and *3)* In the context of other diseases, especially other repeat expansion and neurodegenerative diseases.

## Data Availability

All data produced in the present study are available upon reasonable request to the authors.

https://www.enroll-hd.org

## Acknowledgements

This paper is dedicated to all study participants, care givers, medical professionals, researchers and other support staff who contributed to the unique datasets which enabled this work. Data used in this work was generously provided by the participants in the Enroll-HD study and made available by CHDI Foundation, Inc. Enroll-HD is a global clinical research platform intended to accelerate progress towards therapeutics for Huntington’s disease; core datasets are collected annually on all research participants as part of this multi-center longitudinal observational study. Enroll-HD is sponsored by CHDI Foundation, Inc. Enroll-HD would not be possible without the vital contribution of the research participants and their families. Data used in this work was generously provided by the combined efforts of the Genetic Modifiers of Huntington’s Disease (GeM-HD) Consortium (dbGaP accession number phs000371). NAA is partly supported by a European Research Council Starting Grant (Number: 101041677).

## Appendix Comparison between absolute and relative residual age-at-onset

Let *h* represent the extent of pathology (in arbitrary units) required for the disease to become clinically manifest, and let *t* represent the expected age at clinical onset (in years), assuming a linear increase of pathological burden with time since birth (**Figure A-1**):

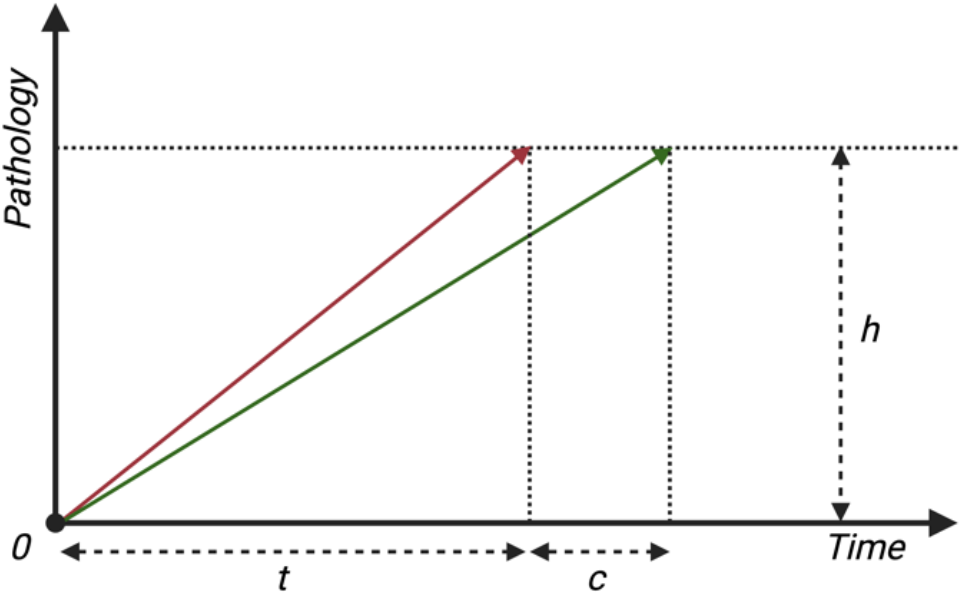

The average rate of *subclinical* disease progression *r* per year could then be defined as:

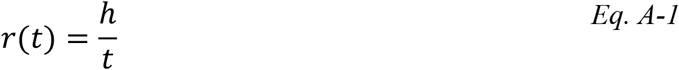

Now suppose that a potential disease modifying genetic variant (or drug) would increase the expected age at onset by *c* years, i.e.:

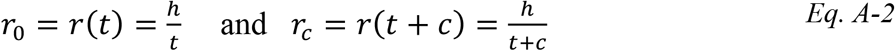

This would have to result in a corresponding decrease in *r* of:

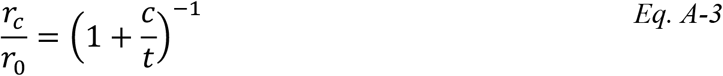

Thus, *Eq. A-3* demonstrates that for smaller values of *t*, the *relative* decrease in the rate of disease progression needs to be greater to ensure a delay in clinical onset by a certain fixed number of years *c* as compared to larger values of *t*, and vice versa. For example, to delay the expected age of onset by 10 years in two premanifest mutation carriers with expected ages of onset of 30 and 40 years, the rate of disease progression would need to be lowered by 25% and 20%, respectively. In contrast, suppose that a potential disease modifying genetic variant (or drug) would change the expected age at onset by a factor *f*, then in the presence of this modifier the rate of disease progression would be given by:

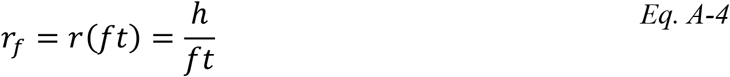

Which will result in a similar *relative* change in the rate of disease progression for different values of expected age at onset (and by extension, for different mutant CAG repeat sizes):

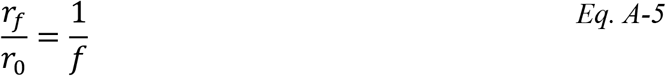

Please note that for the above derivation, it was assumed that pathological burden will increase linearly over time. For completeness, here follows a derivation assuming an exponential increase in pathological burden over time:

Suppose that pathological burden *p* will increase exponentially over time:

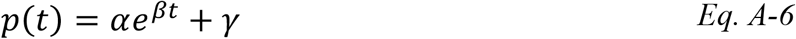

For simplicity, and without loss of generalizability, let’s assume *α* = 1, and *γ* = 0. Now, suppose that a potential disease modifying genetic variant (or drug) would increase the expected age at onset by *c* years, resulting in a change of the growth parameter *β*, i.e.:

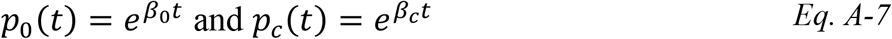

From:

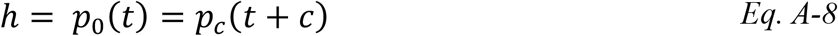

It then follows that:

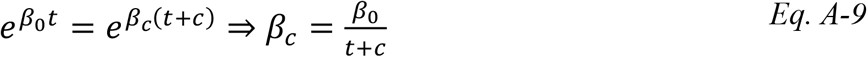

Therefore:

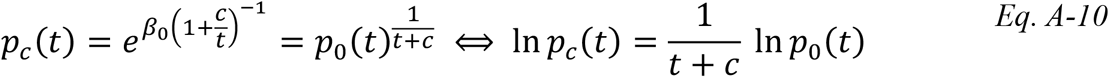

Given that the initial growth parameter *β*_0_ will decrease to a greater extent for smaller values of *t, Eq. A-10* thus demonstrates that also under the assumption of an exponential increase of pathology over time, for smaller values of *t*, the *relative* decrease in the rate of disease progression needs to be greater to ensure a delay in clinical onset by a certain fixed number of years *c* as compared to larger values of *t*, and vice versa. On the other hand, assuming that a potential disease modifying genetic variant (or drug) would change the expected age at onset by a factor *f*, then in the presence of this modifier, for the rate of disease progression the following holds:

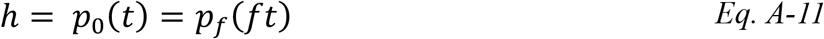

From which it follows that:

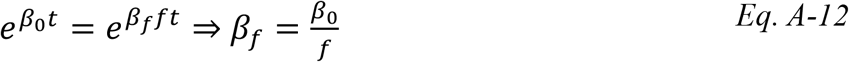

Therefore:

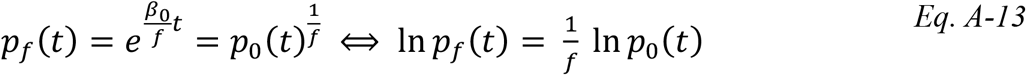

*Eq. A-13* thus shows that when assuming that a modifier will result in a *relative* change of *t*, the growth parameter will simply be scaled to a proportional degree, independent of *t*. This latter is not only biologically more plausible, but also ensures that the effect of the modifier would be easier to compare across individuals with different expected ages of onset (and by implication, also with different mutant CAG repeat sizes).

