## Supplementary Tables and Figures for "Clinical and genetic association studies reveal within-individual variability as an integral part of Huntington disease"

**Supplementary Table 1. Baseline characteristics**

| Sample size (% f) | 7583 (54) | | | | |
|---|---|---|---|---|---|
| | **Mean** | **SD** | **Median** | **25th perc.** | **75th perc.** |
| **Age** | 46.9 | 13.3 | 47 | 37 | 57 |
| **Age of onset** | 48.1 | 12.1 | 48 | 40 | 57 |
| **CAG number** | 43.3 | 3.4 | 43 | 41 | 45 |
| **Composite score** | 12.0 | 4.6 | 12.3 | 8.7 | 15.9 |
| **Total motor score** | 18.9 | 17.4 | 16 | 3 | 31 |
| **Total functional capacity** | 10.9 | 2.7 | 12 | 9 | 13 |
| **SDMT score** | 35.1 | 16.4 | 33 | 22 | 48 |
| **SWRT score** | 74.0 | 24.5 | 74 | 56 | 93 |
| **BMI** | 25.4 | 4.9 | 24.6 | 22 | 27.7 |
| **No. of visits** | 4.9 | 2.0 | 4 | 3 | 6 |

Demographic and clinical characteristics of the study participants at their first study visit. Both Unified Huntington Disease Rating Scale cognitive scores indicate the total number of correct items (in 1 minute).
**Abbreviations:** BMI = body mass index, CAG = cytosine-adenine-guanine, SD = standard deviation, SDMT = symbol digit modalities test, SWRT = Stroop word reading test.

**Supplementary Figure 1. Within-individual clinical variability in Huntington disease increases with larger mutant *HTT* CAG repeat size.** The violin plots depict the association of mutant *HTT* CAG repeat size and the standardized residual within-individual variance for the Unified Huntington Disease Rating Scale composite score, total motor score, total functional capacity, and symbol digit modalities and Stroop word reading test scores, as well as body mass index. Within-individual variability of all clinical measures significantly increased with larger mutant *HTT* CAG repeat size, except for the symbol digit modalities test. For the Stroop word reading test, within-individual variability also increased with CAG repeat size, but the effect tended to level off for larger mutation sizes. The boxes indicate the interquartile ranges around the median (thick black lines), with values deviating more than 1.5 times the interquartile range from the median represented by black dots. For calculating within-individual variability the following approach was used: *1)* a random intercept and slope linear mixed effects model was fitted with sex, age, age squared, mutant CAG repeat size, and an interaction term between age and mutant CAG repeat size as fixed effects, and age as a random effect, and the clinical measures as the dependent variables, *2)* the remaining within individual variability was extracted as the mean of the sum of the squared errors around the fitted random regression lines for each outcome measure, and *3)* a logarithmic transformation (given the right-skewed distribution) and scaling to a standard normal distribution was performed. Comparison among different mutant CAG repeat size grouping were performed using the non-parametric Wilcoxon's rank sum test. WS: within-subject.

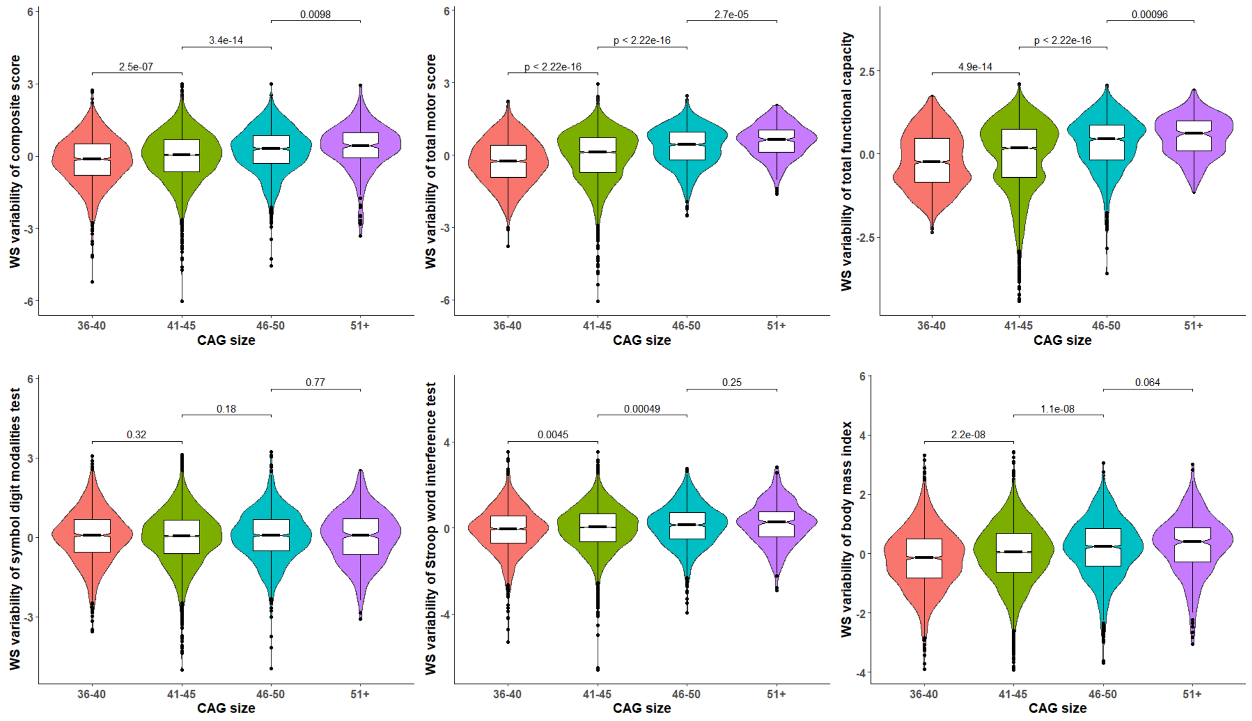

**Supplementary Figure 2. Validation of the analytically predicted variance of action using simulations.** Analytically predicted and simulated variance of action (a measure of phenotypic variability) for a simple Bayesian agent were highly consistent. Both analytically predicted and simulated variance were standardized to a mean of zero and a standard deviation of one for scale-invariant comparisons. See main text for more details.

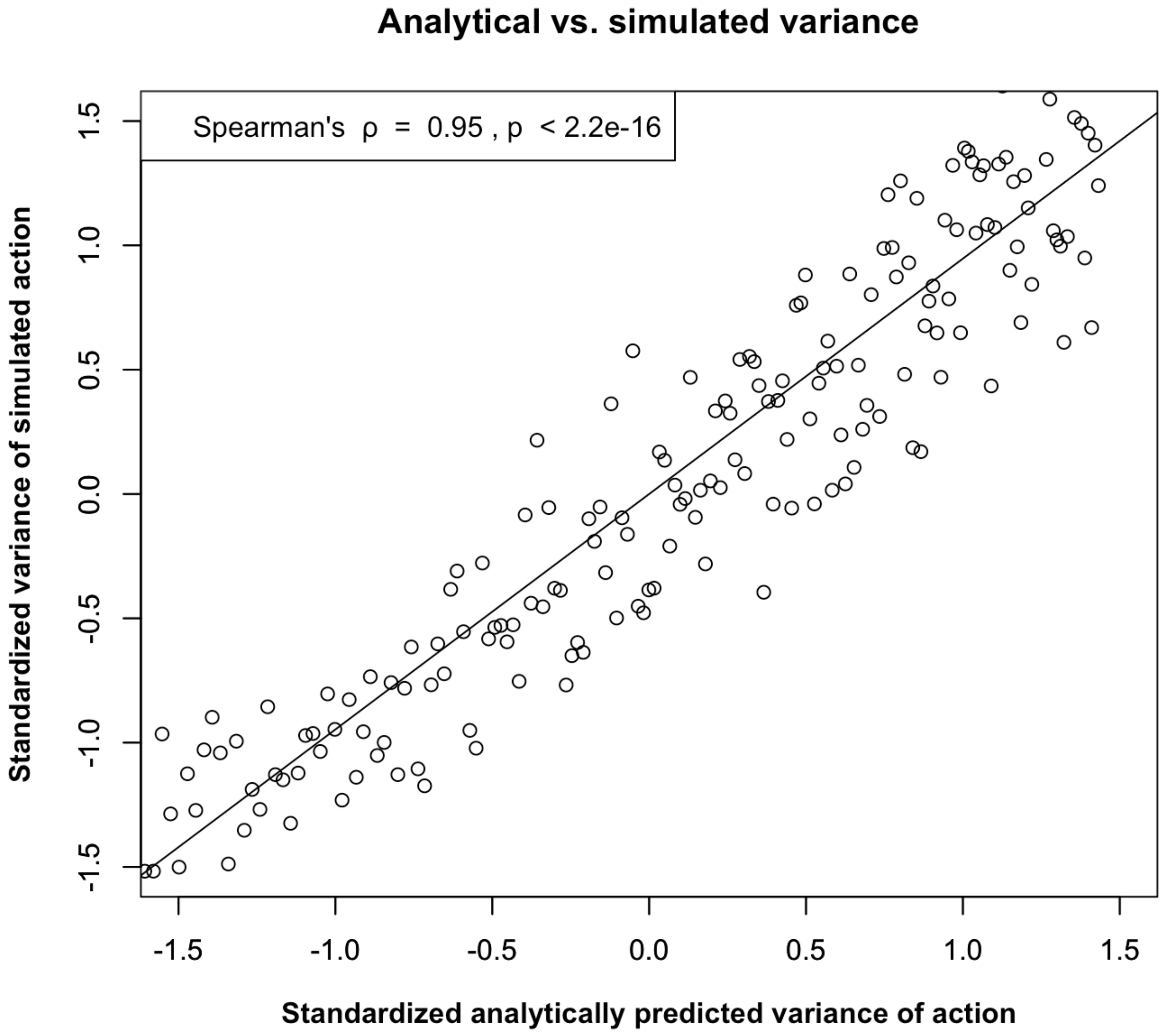

**Supplementary Figure 3. Quantile-quantile plot of the GWAS on body mass index variability.**

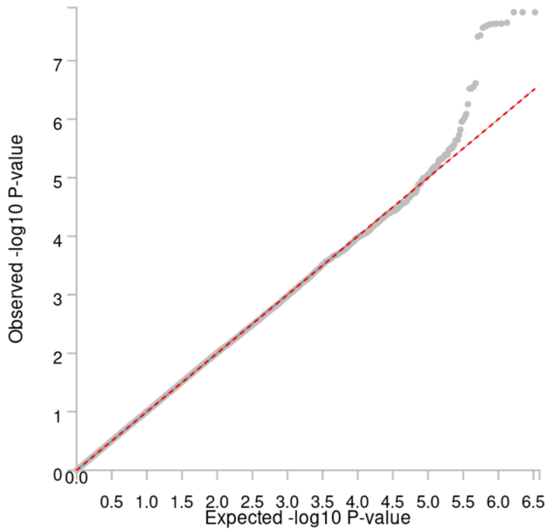

**Supplementary Figure 4. Quantile-quantile plot of the GWAS on mean body mass index.**

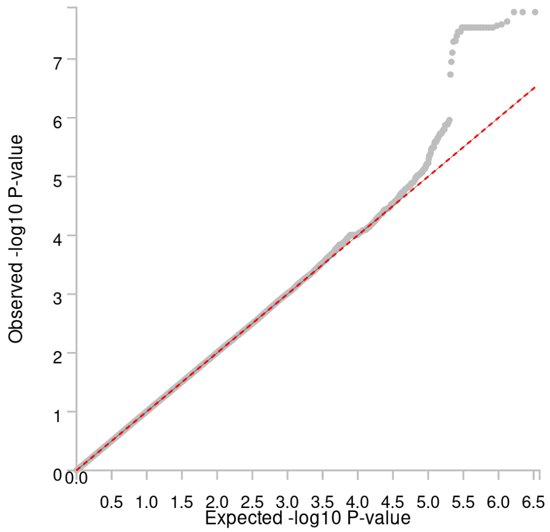

**Supplementary Figure 5. Gene-based Manhattan plot of the GWAS on mean body mass index.** The dashed red line in the Manhattan plot indicates the Bonferroni-corrected gene-based threshold for genome-wide significance (i.e., p = 2.03E-6).

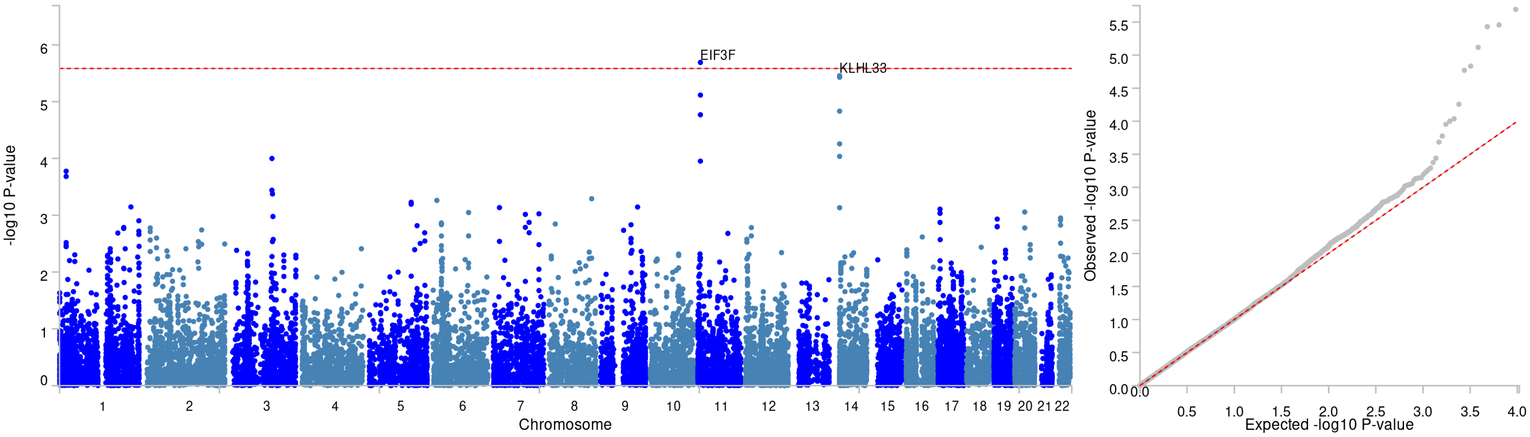

**Supplementary Figure 6. Manhattan and corresponding quantile-quantile plot of the GWAS on variability of Unified Huntington Disease total motor score.** The dashed red line in the Manhattan plot indicates the threshold for suggestive genome-wide significance (i.e., p = 1E-6).

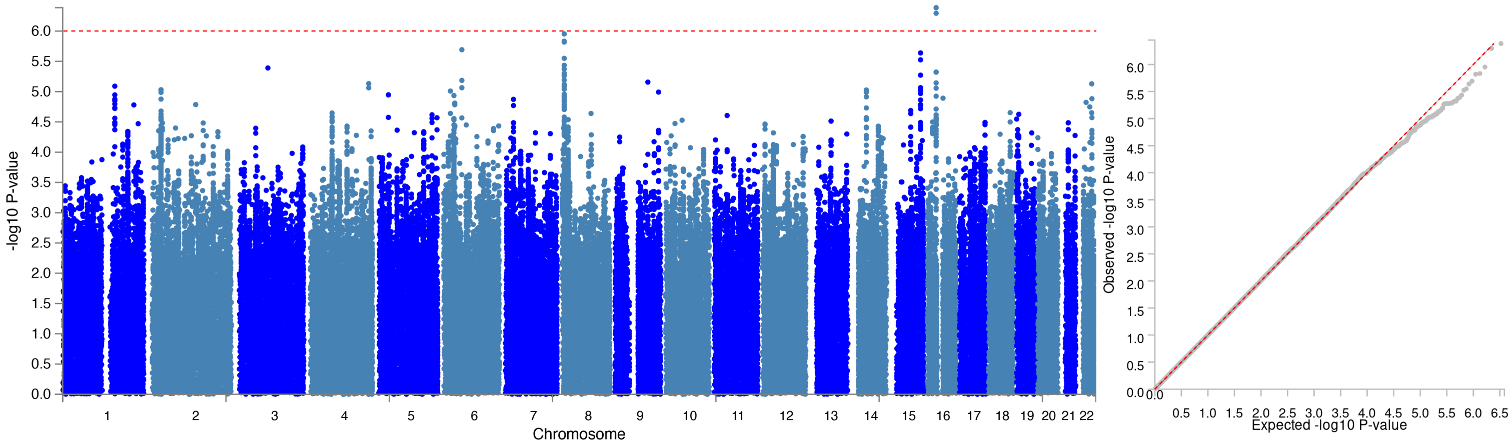

**Supplementary Figure 7. Manhattan and corresponding quantile-quantile plot of the GWAS on variability of Unified Huntington Disease symbol digit modalities test.** The dashed red line in the Manhattan plot indicates the threshold for suggestive genome-wide significance (i.e., p = 1E-6).

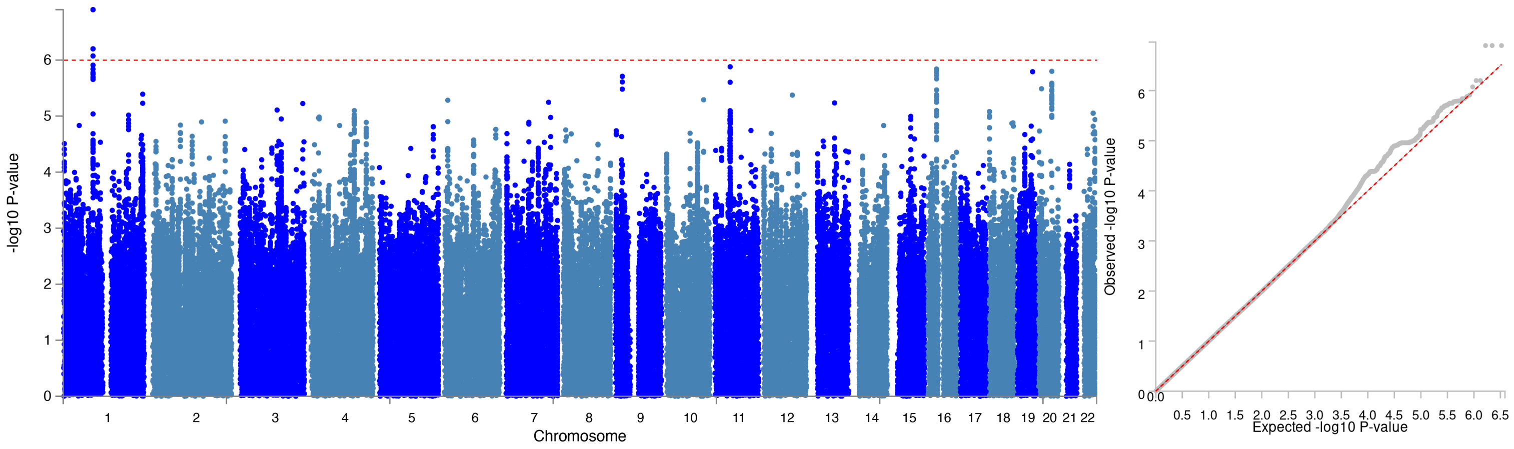

**Supplementary Figure 8. Comparison of the statistical power of using absolute *vs.* relative residual age-at-onset as the GWAS outcome.** Venn diagram comparing the results of the GWAS on residual age-at-onset (RAO, red) *vs.* those of relative RAO (light green), represented by the number (%) of single-nucleotide polymorphisms that passed the threshold for genome-wide significance (i.e., p < 5.0E-8). Overlapping loci (i.e., those detected by both measures) are marked in dark green.

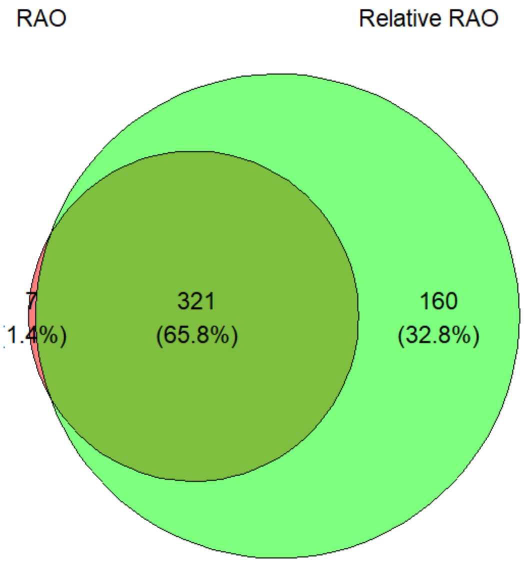
